# Elucidating user behaviours in a digital health surveillance system to correct prevalence estimates

**DOI:** 10.1101/19003715

**Authors:** Dennis Liu, Lewis Mitchell, Robert C. Cope, Sandra J. Carlson, Joshua V. Ross

## Abstract

Estimating seasonal influenza prevalence is of undeniable public health importance, but remains challenging with traditional datasets due to cost and timeliness. Digital epidemiology has the potential to address this challenge, but can introduce sampling biases that are distinct to traditional systems. In online participatory health surveillance systems, the voluntary nature of the data generating process must be considered to address potential biases in estimates. Here we examine user behaviours in one such platform, FluTracking, from 2011 to 2017. We build a Bayesian model to estimate probabilities of an individual reporting in each week, given their past reporting behaviour, and to infer the weekly prevalence of influenza-like-illness (ILI) in Australia. We show that a model that corrects for user behaviour can substantially effect ILI estimates. The model examined here elucidates several factors, such as the status of having ILI and consistency of prior reporting, that are strongly associated with the likelihood of participating in online health surveillance systems. This framework could be applied to other digital participatory health systems where participation is inconsistent and sampling bias may be of concern.

## 1 Introduction

Influenza is a substantial public health concern, with approximately 3-5 million severe cases worldwide each year [1]. There are many challenges in estimating influenza activity and forecasting the spread of influenza, given that disease transmission in the general population is largely unobserved. Traditional influenza surveillance relies on public health system monitoring, such as through hospital admissions, notifications of laboratory confirmed cases, or voluntary reporting from local physicians [2, 3]. However, in these systems, estimates of disease prevalence can be limited by the time taken to collect, collate and publish through public health systems.

Online participatory health surveillance systems attempt to address these challenges by providing a convenient, simple and near real-time platform for self-reporting of symptoms. FluTracking [4] is one such system for monitoring influenza-like-illness (ILI), with a principal aim to contribute to community level ILI surveillance in Australia and New Zealand. Similar platforms exist in the US (Flu Near You [5]) and Europe (Influenzanet [6]).

These platforms often publish estimates of the incidence or prevalence of ILI in the population, usually derived as a proportion of the total number of reports re-ceived that week. These estimates have been found to correlate well with clinical surveillance by public health bodies [7–9], including across different definitions of ILI cases [6].

While systems such as FluTracking show promise in estimating the incidence of ILI in the population, there is evidence that these systems can be effected by variations in user participation [9–12]. Very little is known about how these biases could affect disease prevalence estimates and there is a clear lack of studies that attempt to quantitatively adjust for them. Attempts to correct for these biases have all been based on the removal of data, such as only considering users who frequently participate [6, 7, 13, 14], removing the first report of a user [12], or by creating noise filtering algorithms that minimise sudden departures from sentinel data due to changes in participation [15]. While not unreasonable, these methods do not give insight into user behaviour, and do not examine the effect of their corrections on the estimates they examine. Some examinations of the heterogeneity of users and their behaviours have been conducted, such as inferring significant predictors for and classifying users on their participation levels [16, 17], and examining the demographic representativeness of the participating population [14, 17, 18]. However, user behaviour has not been analysed within a systematic, statistical modelling framework, in particular one which can simultaneously correct disease prevalence estimates. This work addresses this gap by using information about how individuals behave with regard to survey completion to inform our estimate of the weekly prevalence of ILI. While the scope of this work is limited to Australia, the analysis is generalisable to other types of voluntary, web-based surveillance in other locations and settings, of which there are numerous.

### FluTracking Data

Participants can register in FluTracking at any stage of the year, and upon registration, they provide various demographic details about themselves, including age, gender, and postcode. Participants can optionally register and report on behalf of others in their household. Those participants who are submitting reports, on behalf of themselves and/or their household, will henceforth be referred to as ‘masters’, while any individual observed, master or not, will continue to be referred to generally as ‘participants’. After registering, masters are sent an email on the Monday of each week during the influenza season to respond to an online questionnaire about the presence of fever or cough that they or their household members may have experienced in the week prior, and whether they have received the influenza vaccine this year.

Surveys can be submitted for up to 5 weeks from their first reminder. Note that FluTracking often publishes estimates of ILI incidence to examine the spread of new cases of ILI [19]. In this study, we will focus on estimates of prevalence in order to determine if user behaviour is influenced by ILI status.

If a participant reports both a fever and a cough, follow up questions are revealed, such as enquiring about any health-seeking behaviour taken or if a sore throat was experienced. In this study, we define an ILI case as a survey response with a fever and a cough, which closely resembles the World Health Organization surveillance case definition [20].

Note that a report, and therefore an observation of ILI status in a household, is generated by the action of the master, and so it is the report of the master and subsequent reports on behalf of the rest of the household that is the outcome of interest. Also note that in order to build a framework for near real-time prediction, we have chosen to define a report submitted less than 7 days after the initial request as an ‘on-time’ report and not a ‘late’ report. This is the interval of time before the subsequent request for the next survey is sent. However, if symptoms are submitted in a late report, this information is not excluded, but used in the derivation of predictor variables for subsequent survey weeks, irrespective of submission date.

FluTracking operates between May and October every year in the winter season in the Southern Hemisphere. We examine all Australian reports submitted between 2011 and 2017, totalling 3,459,339 unique reports from 30,564 households and 52,773 participants. Of these reports, 352,287 (approx. 10%) are late reports.

Using the registration date of individuals, we can infer the weeks in which household reports were missed and never submitted as survey weeks without a report after their registration date for each season. This includes another 496,175 missed surveys.

### The Model

The conventional estimate in the literature is given by the total number of individuals that have reported with ILI, divided by the number of on-time reports in the given week. We improve this by developing a framework to adjust ILI prevalence estimates by correcting for user behaviour, and construct a model to predict the probability of a user reporting in a given week, based on their prior behaviour and demography (Figure 1). An individual household *i* in a given week *w* will receive a survey request to participate, and will then proceed to either report and provide information on their symptoms, or not report. Given that they report, and their symptoms are therefore known, they will either report on-time, or report late. We will compare the following two estimates of the prevalence of ILI:

**Figure 1:**
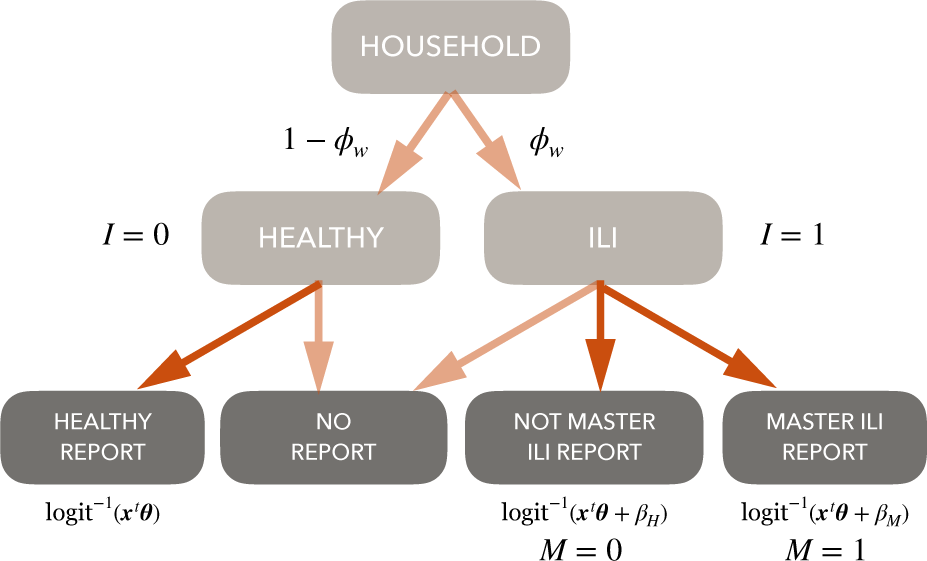
Visualisation of model of user outcomes for a household *i* in week *w*, where the probability of reporting given their health status is modelled with the link function listed below respective compartment. The probability of having at least one household member with ILI is modelled with *ϕ*_*w*_ (Equation 2).

- the naïve estimate (an extension of the conventional estimate); and,
- a behaviour corrected estimate, (our new framework).

Our naïve estimate extends the conventional by considering a Bayesian perspective on the estimate, which respects the same mode value as the conventional estimate. The behaviour corrected estimate is inferred from a framework that incorporates an observation process in the model, whereas the naïve model does not. Both estimates assume the number of individuals with ILI in the population is binomially distributed.

## Materials and Methods

### Naïve Model

The conventional estimate takes the total number of individuals that have reported with ILI this week *X*_*w*_ and divides this by the number of on-time reports this week 𝒩_*w*_. If we consider that each individual in the population has an equal probability 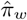 of having ILI in week *w*, then *X*_*w*_ can be modelled with a binomial likelihood with probability 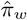 and trials 𝒩_*w*_. Note that this interpretation considers 𝒩_*w*_ as fixed and not a random variable. Given a uniform prior Beta(1, 1) and a binomial likelihood, the posterior distribution of 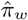 is then Beta(*X*_*w*_ + 1, 𝒩_*w*_ − *X*_*w*_ + 1). This results in the mode of the posterior distribution of 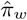 as 𝒩_*w*_ */ X*_*w*_, conventional point estimate often used in the literature (See [6, 7, 15, 21]). We define this posterior distribution of 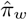 as the naïve estimate, which is an estimate for the prevalence of ILI in the population, without consideration of user behaviours. In the work presented here, we did not split the cohort by vaccination status and model estimates for vaccinated and unvaccinated prevalence. Models using a split cohort with separate parameters for vaccinated and unvaccinated prevalence were examined, with only marginal differences in the corresponding parameters, typically near the peak ILI week. As such, we have simplified the model and reduced the number of parameters inferred. While it can be tempting to examine the difference between the vaccinated and unvaccinated estimates as a measure of influenza vaccine effectiveness, in this study we examine cases of ILI, not influenza. Any examination of vaccine effectiveness will require further modelling.

### Behaviour Model

We construct a model (Figure 1) that accounts for user behaviour and informs an estimate for the prevalence of ILI in the population, for vaccinated and unvaccinated individuals. For a household *i* in week *w*, let *Y* be a binary indicator of an on-time report submitted by the master of the household, *I* be a binary indicator of at least one participant of the household having ILI, and *M* be a binary indicator of the master of the household having ILI.

For every household in every week, there can be one of four outcomes

- A household submits a report on-time, and no participants report having ILI (*Y* = 1; *I* = 0),
- A household submits a report on-time, and at least one member that is not the master reports having ILI, (*Y* = 1; *I* = 1;*M* = 0),
- A household submits a report on-time, and the master reports having ILI, (*Y* = 1; *I* = 1;*M* = 1),
- A household does not submit a report on-time (*Y* = 0).

Logistic regression can be used to estimate a household’s probability of reporting *p*, where for some set of predictors ***x***_*i,w*_ and parameters ***θ***, *β*_*H*_ and *β*_*M*_. The link function is defined as

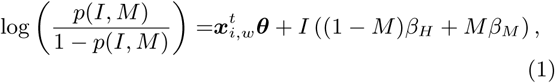

where

- ***x***_*i,w*_ is the vector of predictors for reporting,
- ***θ*** is the parameter vector of regression coefficients,
- *β*_*H*_ is the parameter for the change in reporting behaviour due to the household having at least one member with ILI, but not the master, and
- *β*_*M*_ is the parameter for the change in reporting behaviour due to the master having ILI.

Assume each household member has an independent probability *π*_*w*_ of having ILI in week *w*, then we can define *ϕ*_*w*_ to be the probability that a household of size *n*_*i*_ has at least one individual with ILI in a given week *w*. The probability *ϕ*_*w*_ can then be modelled as

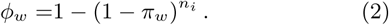

The expression of the probability of the master having ILI *M* and reporting *Y* can be seen below. The other three possible outcomes for a given household in a given week can be similarly derived, and is not presented.

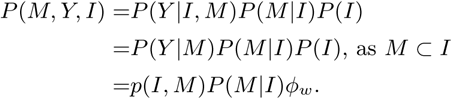

Defining *ζ* = *P* (*M*|*I*), an expression for *ζ* is:

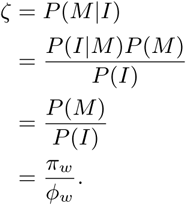

The likelihood ℒ is then the product of the likelihood of each household *i* in each week *w* in the training set

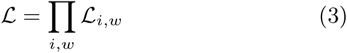

Where

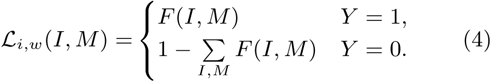

and

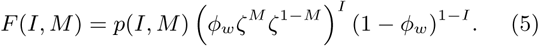

### Model Fitting

We use a Bayesian framework to estimate the posterior distribution of the parameters ***θ, β*** = (*β*_*H*_, *β*_*M*_), and ***π***, conditional on the data ***X*** and ***y***, via Bayes’ rule:

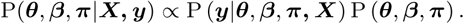

Prior distributions for the parameters were:

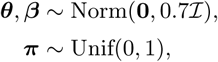

where **0** is the zero vector and ℐ is the identity matrix.

The variance of the prior distributions for ***θ*** and ***β*** were chosen to provide a near uniform prior density for the probability of reporting and reporting on-time, given the distribution of the training set predictors. Figure S1 in the Supplementary Information shows the prior distribution transformed from the log odds scale to the probability scale, after multiplication with samples from the training set predictors. Results were not sensitive to this choice of variance.

All predictors ***x***_*i,w*_ were scaled to be between 0 and 1, to allow for simplicity in comparing the predictive strength of each parameter. The set of predictors used for regression were derived from variables listed in Table 1, and the complete set of predictors used can be seen in Table S1.

**Table 1:**
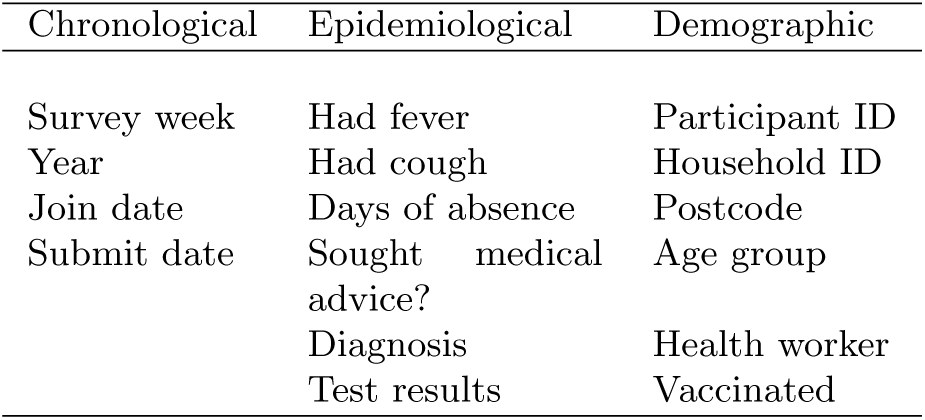
Variables from the dataset used in constructing predictors.

To estimate the posterior distribution of the parameters of the model, we used Hamiltonian Monte Carlo (HMC) implemented in the software package Stan (version 2.18) [22]. HMC is particularly effective over other Markov chain Monte Carlo methods, such as Random Walk Metropolis-Hastings, in high dimensional parameter spaces such as in this analysis, and the Stan implementation uses adaptive tuning of algorithm parameters, reducing the need to tune the inference algorithm [23].

For model training and inference of results, 80% of the households in each year were used in the training set for the model, and the remainder left in a test set for cross-validation of model predictions. For each season examined, the first week of reports was used in determining the prior behaviours of users and used in calculating predictor values, but were not used to infer parameters of the model, as predictors involving prior behaviours could not be well defined in the first week.

### Data Access

Reproduction of this study would require individual level data, which is precluded by ethics approval (The University of Adelaide Human Research Ethics Committee (HREC) H-2017-131). Anonymised data may still allow the possibility of identification of individuals, given the rich set of features and the small numbers in some post-codes.

## Results

### Comparison to Naïve Estimate

Without correcting for user behaviour, our results show that naïve estimates of ILI prevalence are biased and over-estimate the actual prevalence. The distribution of the naïve estimates have nearly all their probability density higher than the distribution of the behaviour corrected estimates for the ILI prevalence (Figure 2). This bias was found to be greatest when prevalence of ILI was greatest, and implies that users may be more likely to report when they have ILI. This trend was observed for every year of analysis (see Supplementary Information). Figure 2 shows the results from training the model on the year 2017.

**Figure 2:**
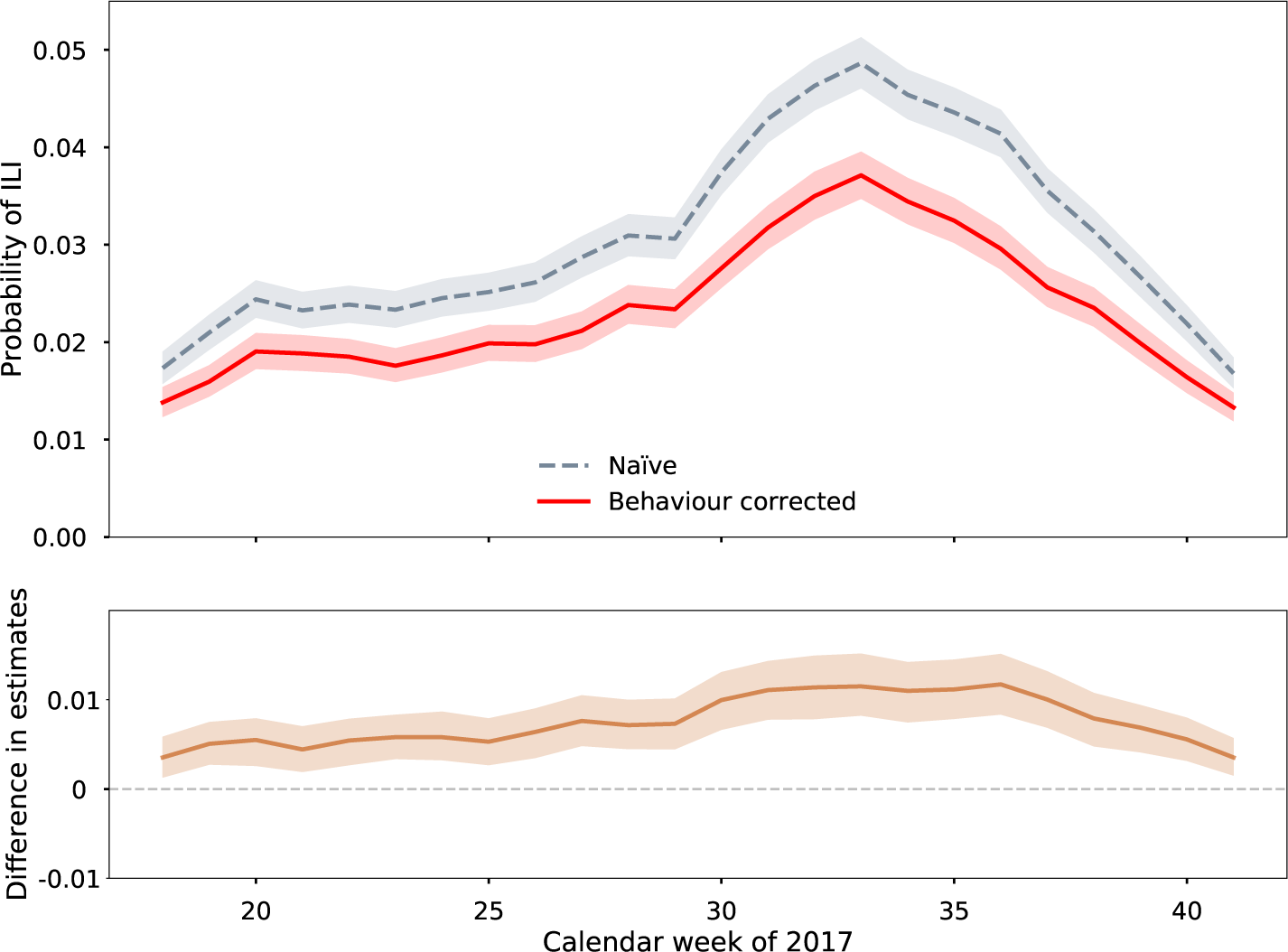
Comparison of behaviour corrected posterior estimate of the prevalence of ILI in the population (solid) to the naïve estimate (dashed) for the winter of 2017, and the difference between the two distributions over time. Lines represent the median of the distribution and shaded regions are 95% credible intervals of the posterior distributions.

### Reporting Behaviour

Predictors of reporting behaviour were determined from chronological, epidemiological and demographic data (Table 1). Given a consistent identification number for each individual and each household, predictors such as the week of the survey, whether the individual reported having a single symptom previously this year, and whether an individual is reporting on behalf of others can be determined. A full list of predictors used and their explanations can be found in Table S1.

Unsurprisingly, the greatest predictor for reporting behaviour was found to be the proportion of reports submitted on-time in the year so far for an individual, with all posterior samples of log odds ratio greater than 4 (see Figure S10). Here we will present predictors of interest, and the marginal densities of all predictors are presented in the Supplementary Information.

A comparison of marginal posterior densities of regression coefficients for predicting the probability of reporting (*p*) can be seen in Figure 3. Those reporting on behalf of others were not found to be much more likely to report, inconsistent with analysis in other studies [16, 17]. These studies did not consider the past reporting behaviour of users in their analysis, and this may have confounded this predictor in previous work. Individuals who have reported being vaccinated for this season were also found to be more likely to report in any given week. Individuals who had reported having a symptom in previous weeks were much more likely to submit a report. However, the increasing number of weeks since the symptom occurred was a strong predictor for not submitting a report. Given no other differences, the model then predicts an increase in probability for an individual to report when they have reported having a symptom, with the increase decaying over time as the number of weeks since reporting the symptom increases. Figure 3 also shows that there is a large increase in the log odds of reporting when the household, but not the master, has ILI. However, when the master has ILI, there is a substantially larger increase in log odds or reporting. This would indicate that the status of having ILI in proximity, and in particular personally experienced, is a strong predictor reporting and participating in FluTracking. The posterior densities are remarkably consistent across the years, with the direction of the effect of the predictor consistent in all years, with the exception of those with small effects.

**Figure 3:**
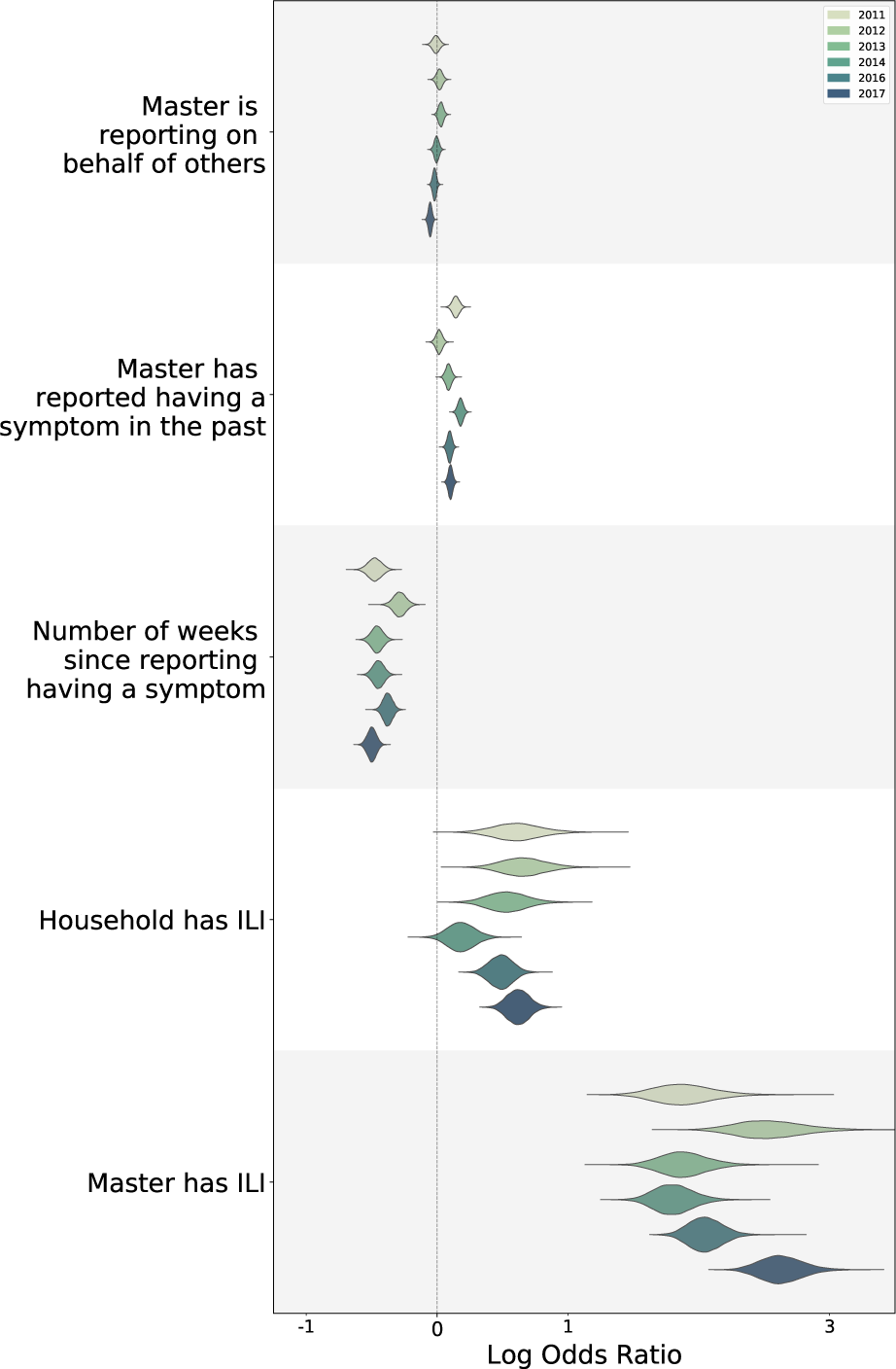
The marginal posterior densities of the log odds ratio of certain regression coefficients for predicting the probability of an individual reporting on-time across all years examined. For the complete set of regression coefficients, see the Supplementary Information.

The current week was also used as a predictor for the probability a user will report, and their marginal probabilities of reporting declined sharply in the first third of the season, before a more shallow decline as the season continues (Figure 4). This could be explained by user fatigue, where dedication to participating in a voluntary system wanes through the year. This trend was also seen in every year studied (see Supplementary Information).

**Figure 4:**
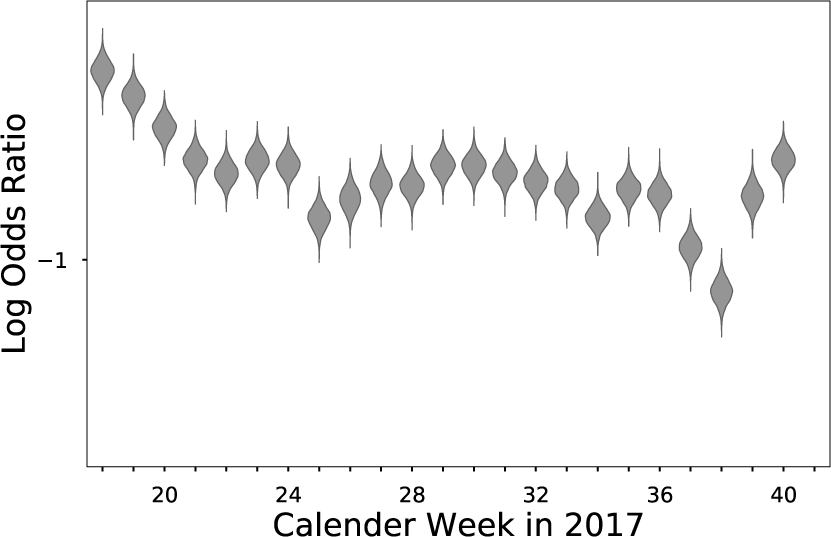
The marginal posterior densities of the log odds ratio of chronological regression coefficients for predicting the probability of an individual reporting in a given week of 2017. For all other years, please see Supplementary Information.

### Model Validation

To validate the model, 20% of the households in each year were excluded from training the model and kept as a test set. Model predictions were compared to observations in the test set by taking 1000 samples of the parameters from the posterior distribution, and simulating outcomes from the model for each sample and each household in the test set. The simulated data classified each household in each week into one of the four outcomes (Figure 1).

The proportion of reporting households with at least one participant with ILI, and the participation rate of households of the simulated data, were compared to observations from the test set. The proportion of reporting households with at least one participant with ILI was calculated by dividing the number of households reporting on-time with ILI by the total number of households reporting on-time. The participation rate of households is determined by dividing the number of households reporting by the total number of households registered who have submitted at least one report in that year. Both summary statistics from the data were similar to simulations from the model on the test set using samples from the posterior, and these comparisons can be seen in the Supplementary Information Figures S13 to S19. The simulated data from the model matches well to the actual observations from the test set and appears to retain the autocorrelation or time dependence of the actual test outcomes, without time dependence being explicitly included in the model. As the model predictions correspond well to outcomes observed in the test set in both summary statistics, the model shows no indication of bias of observable outcomes at a population level.

## Discussion

Online participatory health surveillance systems strive to provide near real-time estimates of disease prevalence, and yield complementary insights to traditional public health systems. However, the voluntary nature of these systems, and the reliance on user participation, need to be considered.

In this work we find that the presence of ILI in the household, as well as other demographic factors, impacts the probability a user will submit a report. At a population level in FluTracking, this results in overestimation of the prevalence of ILI in the population when using the naïve estimate. This difference is greatest near the peak prevalence of ILI. This may be due to users being triggered to report by their symptoms, and being more likely to report in voluntary health surveillance systems when directly affected by the illness, through their household, or more strongly by personal experience.

Participation in FluTracking, derived as the number of on-time reports divided by the number of registered users reporting at least one in a given year, generally ranges between 70% and 90% for any given year and any given week. This is much higher than rates observed in Influenzanet and Flu Near You, where previous studies have shown that 70% and 35% of users respectively submit more than 3 reports in a season [16, 17]. This is despite FluTracking having proportionally much higher numbers of participants than the European and US systems, relative to the respective populations of these regions. While the variance of participation over time has not been examined in the other systems, correcting for user behaviour would potentially be even more critical than observed here.

The estimates of the prevalence of ILI were not modelled with influenza vaccination status incorporated. As mentioned earlier, models with vaccination status included were considered, but are not presented here. Models which produce simple ILI prevalence estimates, as examined here, can not be used as a measure of influenza vaccine effectiveness. The vaccine is not targeted to ILI, and the changes in prevalence over time does not consider the population size, or the number of cases of influenza reduced in a season. For these reasons, vaccination status was not incorporated into the model for prevalence estimates.

However, there exists the potential to construct a measure of vaccine effectiveness by comparing the two groups. The conventional metric of test-negative cases [24] does not extend simply to the behaviour corrected estimates presented here. However, with the potential to correct for potential bias in the data and reporting behaviour across different years, further studies may help inform vaccine effectiveness estimates in near real time.

Whilst past behaviours and illness status were used to predict user participation within the season, this study did not attempt to train the model across seasons. Training the model across every season would allow for users’ behaviour in past years to inform behaviour in later seasons. However, the computational difficulty in substantially increasing the size of the training data, and the number of parameters involved, remains a challenge left for further work.

Predictors for user behaviour have only been taken from within the FluTracking data set. Social media and news coverage [25] and public awareness [26] are some examples where external factors can influence behaviour during an epidemic. Inclusion of these factors in the model may further improve estimates of disease prevalence where user behaviour may be significant.

This analysis assumes each individual has an equal probability of having ILI in a given week. Incorporating a spatial mechanistic model, particularly given the post-code and demographic information in the data set may enable further insights into the mechanisms that drive the transmission of influenza and ILI in Australia. Recent analysis has shown the onset of influenza epidemics is largely synchronised across regional areas [27].

With this model we have shown that user behaviour can have a significant effect on disease prevalence estimates drawn from the data. The framework developed herein, which elucidates drivers of user reporting in voluntary health surveillance systems and improves estimates of disease prevalence, should prove useful as digital data streams burgeon in epidemiology.

## Data Availability

Reproduction of this study would require individual level data, which is precluded by ethics approval (The University of Adelaide Human Research Ethics Committee (HREC) H-2017-131). Results, simulations used for validation and code is linked below.

https://tdennisliu.github.io/publications/FTBehaviour/Supplementary_Data.zip

## Acknowledgements

We would like to acknowledge the FluTracking team for their support, and all the FluTracking users for participating. This work was supported with supercomputing resources provided by the Phoenix HPC service at the University of Adelaide. Funding and support was provided by the Data To Decisions Collaborative Research Centre (D2D CRC), the Australian Research Council Centre of Excellence for Mathematical and Statistical Frontiers (ACEMS), the National Health and Medical Research Council (NHMRC) Centre of Research Excellence in Policy Relevant Infectious diseases Simulation and Mathematical Modelling (PRISM^2^), and an Australian Government Research Training Program (RTP) Scholarship.

## Supplementary Information

## Methods

Table S.1 presents all predictors used in the analysis. Figure S.1 depicts the choice of prior distribution for the regression coefficients.

## Results

All figures are presented in reverse chronological order, as the more recent years have the most directly comparable populations of participants to 2017, the year that was primarily discussed in the main paper. Figures S.2 to S.7 compare the naïve estimate to model posterior distributions of ILI prevalence in vaccinated and unvaccinated individuals across all years studied. 2017 results are presented in the main article. Figures S.8 to S.12 show the marginal posterior densities of the log odds ratio of all regression coefficients for predicting the probability of an individual reporting on-time across all years examined. Figure S.11 shows some of the bivariate kernel densities of certain parameters of the posterior, displaying the lack of correlation between parameters in the posterior. Figures S.13 to S.19 compare the model predictions against summary statistics of the test sets across all years examined.

**Table S1:**
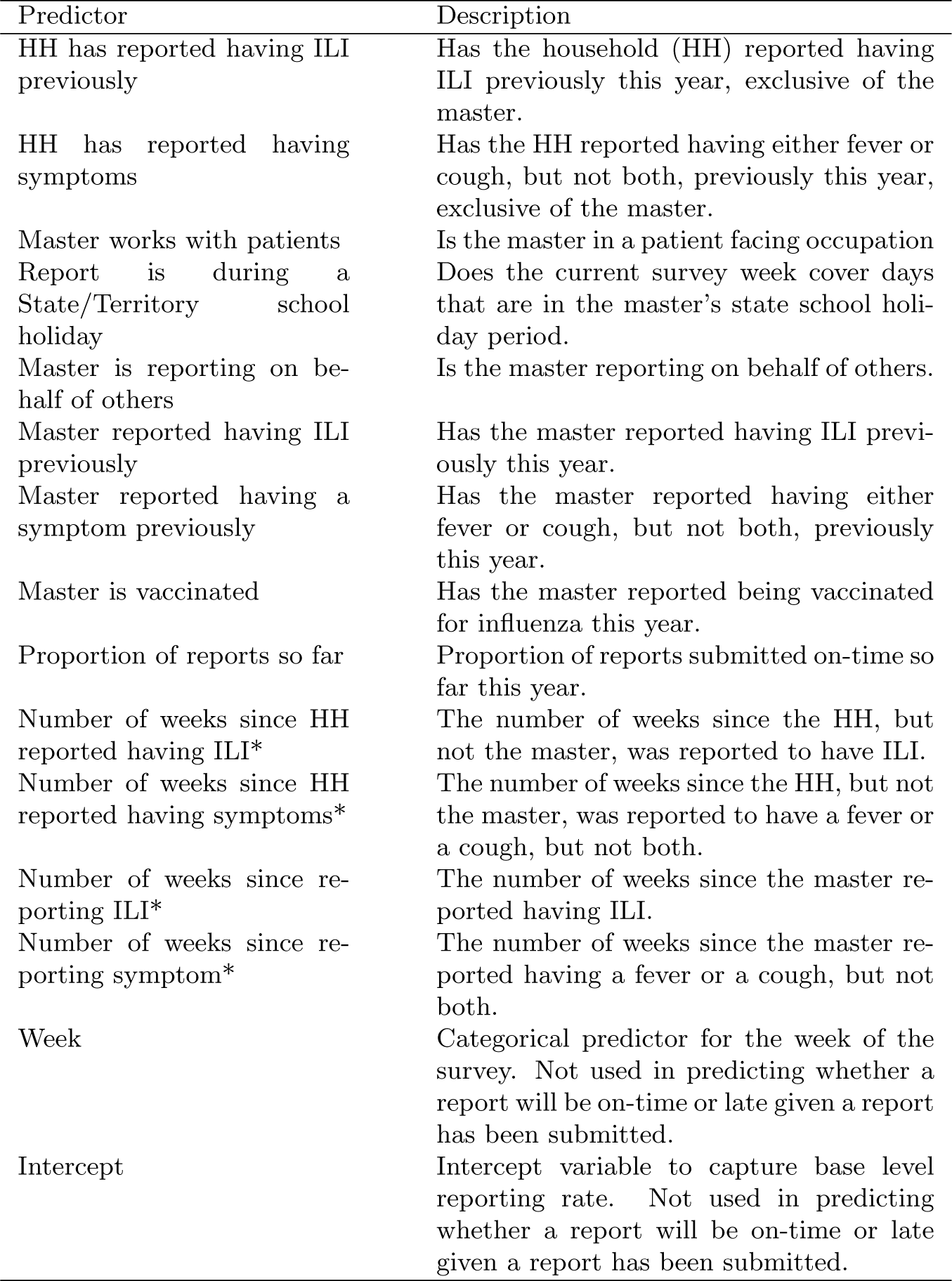
List of predictor variables used and their definitions. Asterisks indicate variables in which the value is divided by the number of weeks in the season to scale the predictor between 0 and 1. School holidays breaks are sourced from Australian government sources [28].

**Figure S.1:**
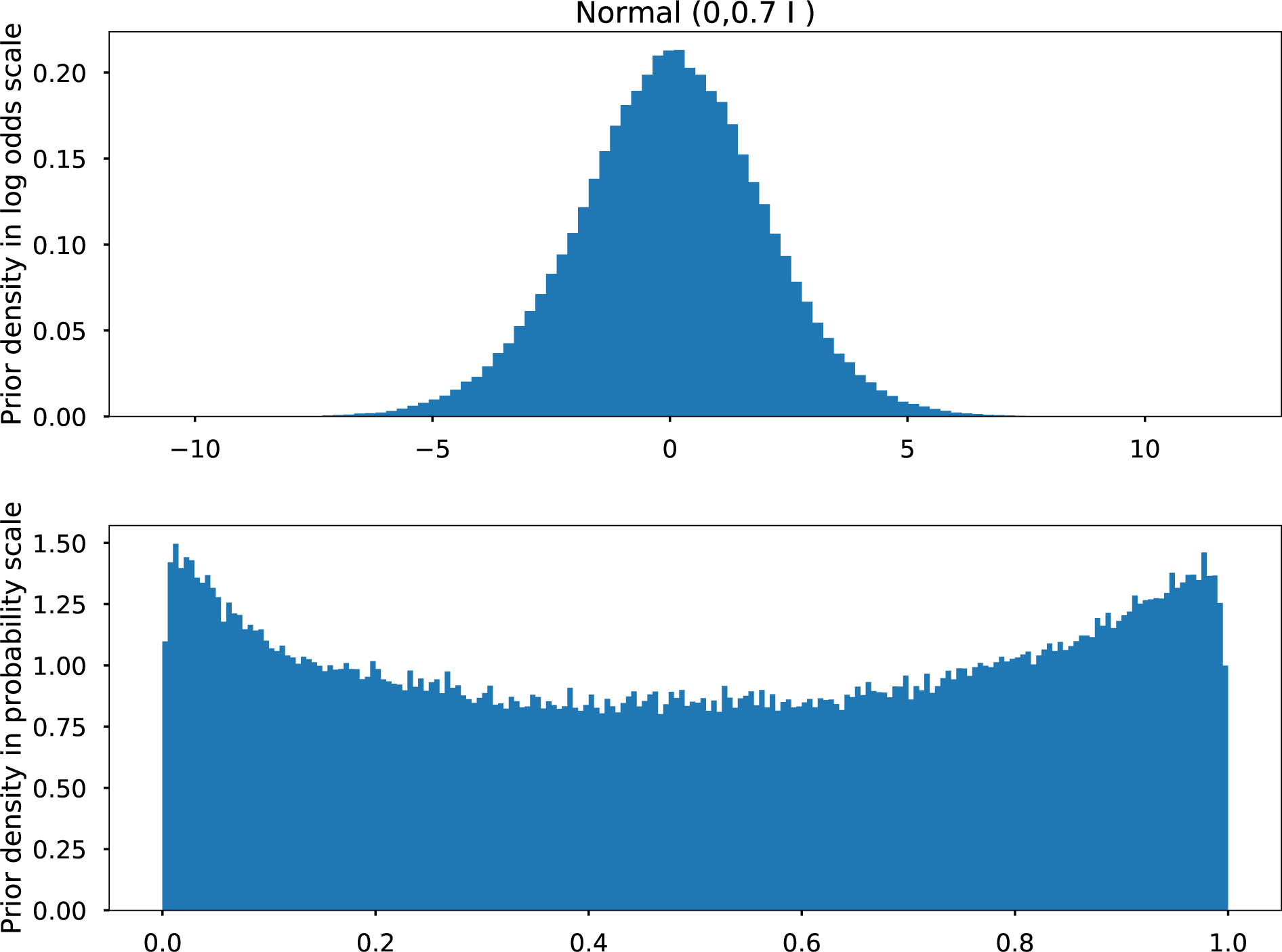
The prior distribution of the regression coefficients transformed from the log odds scale to the probability scale via the logit function, after multiplication with 1000 samples from the training set predictors of 2017. The covariance matrix 0.7 ℐ was chosen for the model as it was somewhat uniform across the probability scale, with some skewness towards the boundaries.

**Figure S.2:**
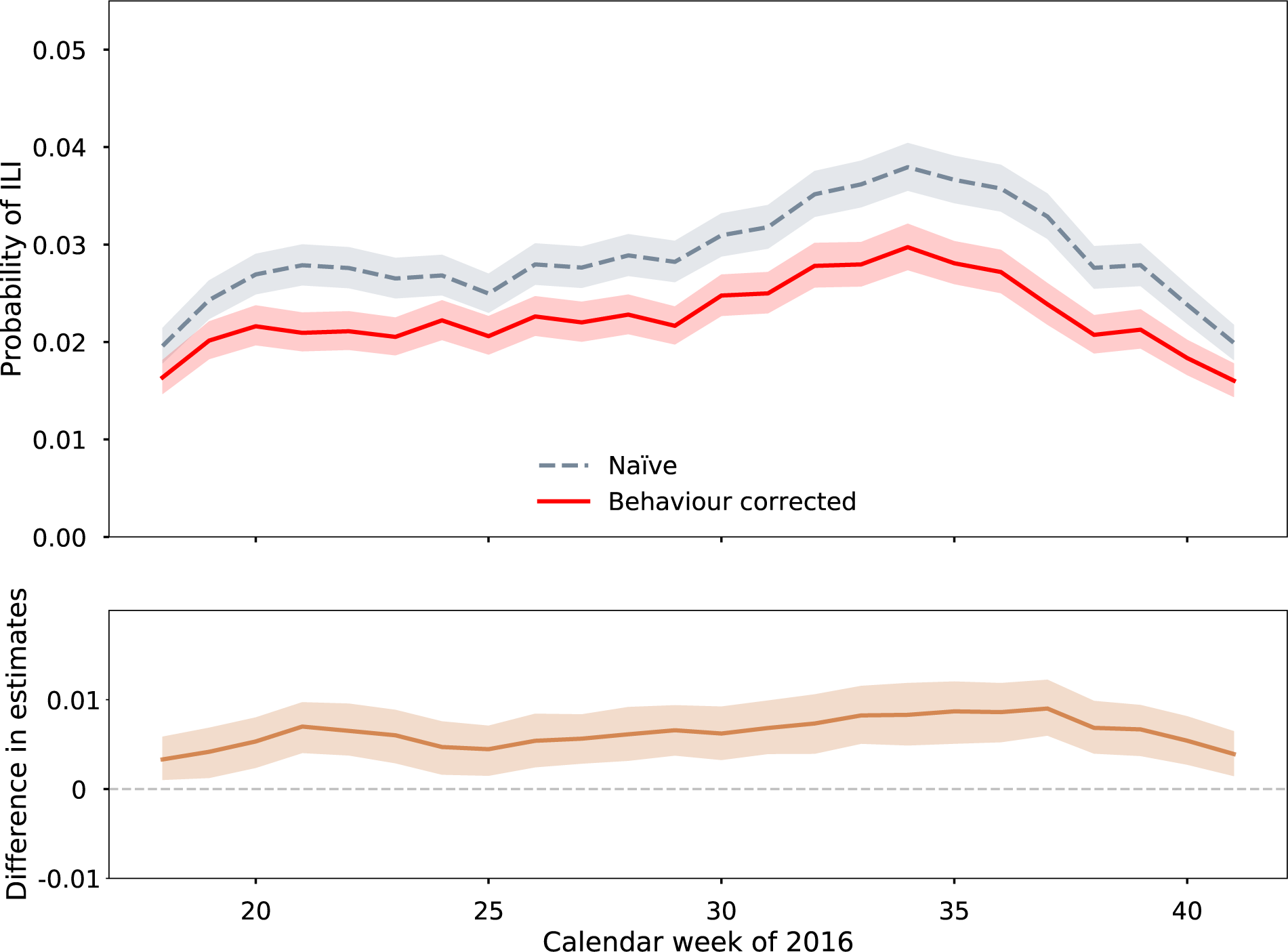
Comparison of model posterior estimate of the prevalence rate of ILI in the population to the naïve estimate for the winter of 2016, and the difference between the two distributions over time. Lines represent the median of the distribution and shaded regions are 95% credible intervals of the posterior distributions.

**Figure S.3:**
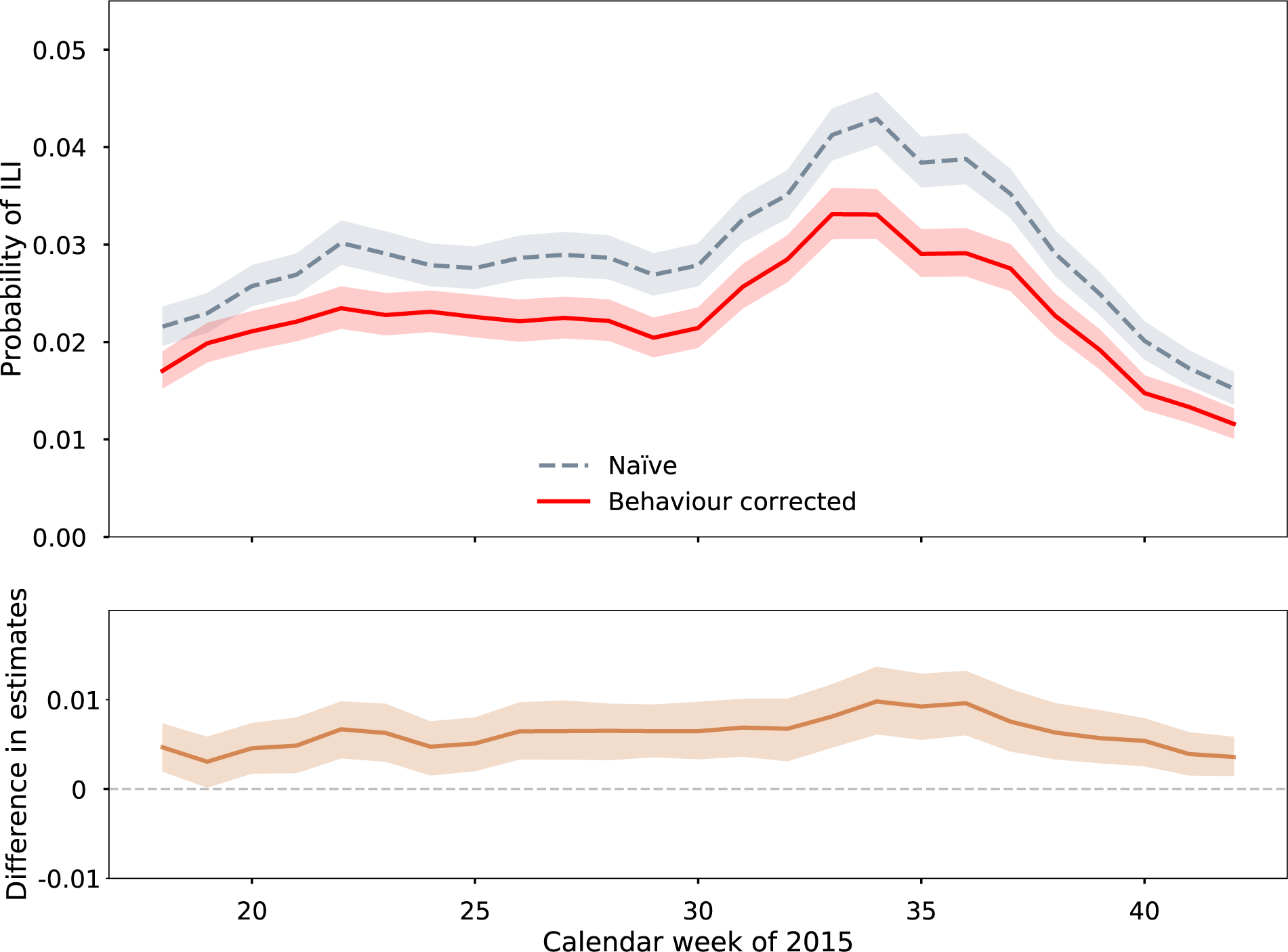
Comparison of model posterior estimate of the prevalence rate of ILI in the population to the naïve estimate for the winter of 2015, and the difference between the two distributions over time. Lines represent the median of the distribution and shaded regions are 95% credible intervals of the posterior distributions.

**Figure S.4:**
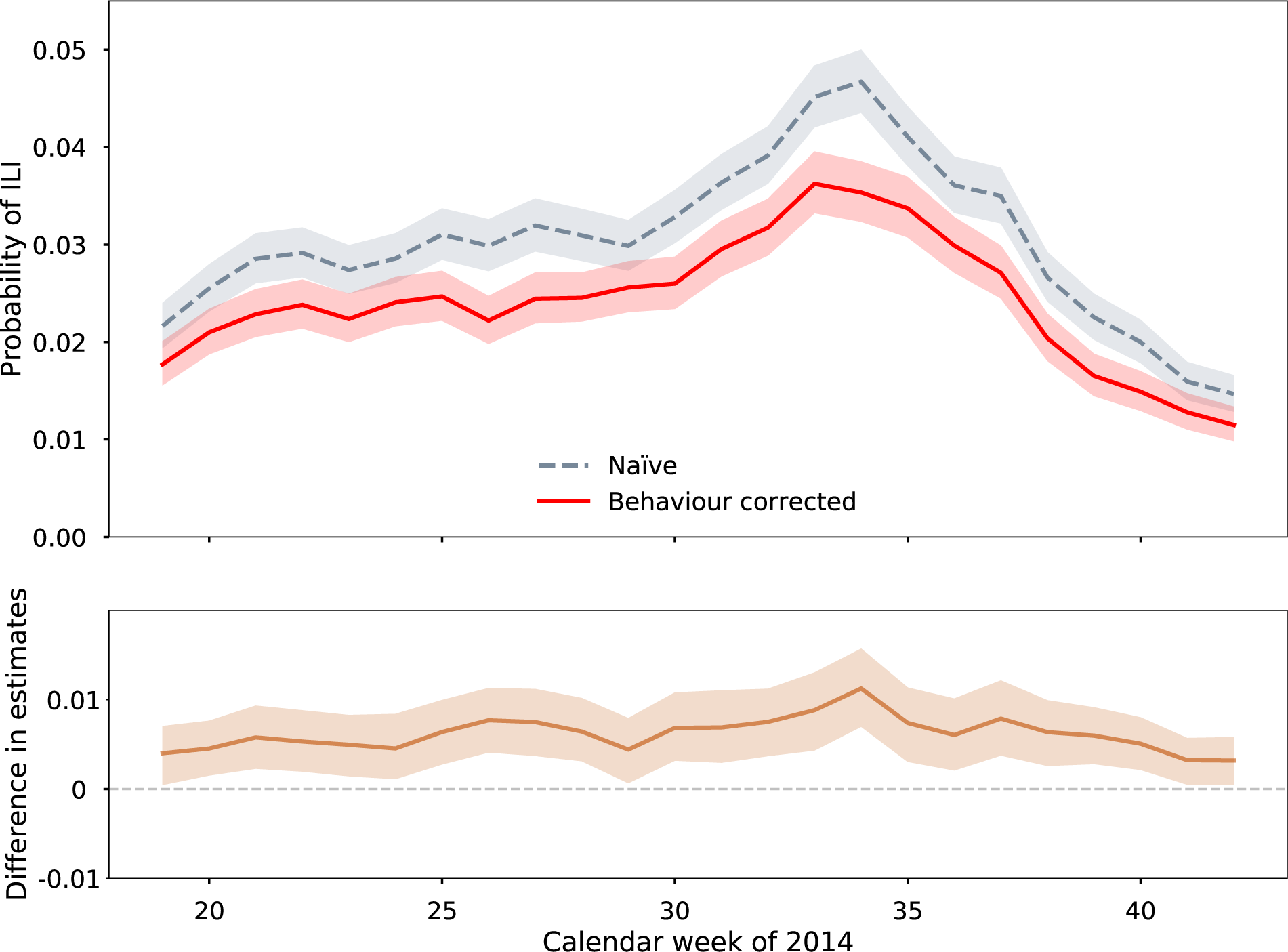
Comparison of model posterior estimate of the prevalence rate of ILI in the population to the naïve estimate for the winter of 2014, and the difference between the two distributions over time. Lines represent the median of the distribution and shaded regions are 95% credible intervals of the posterior distributions.

**Figure S.5:**
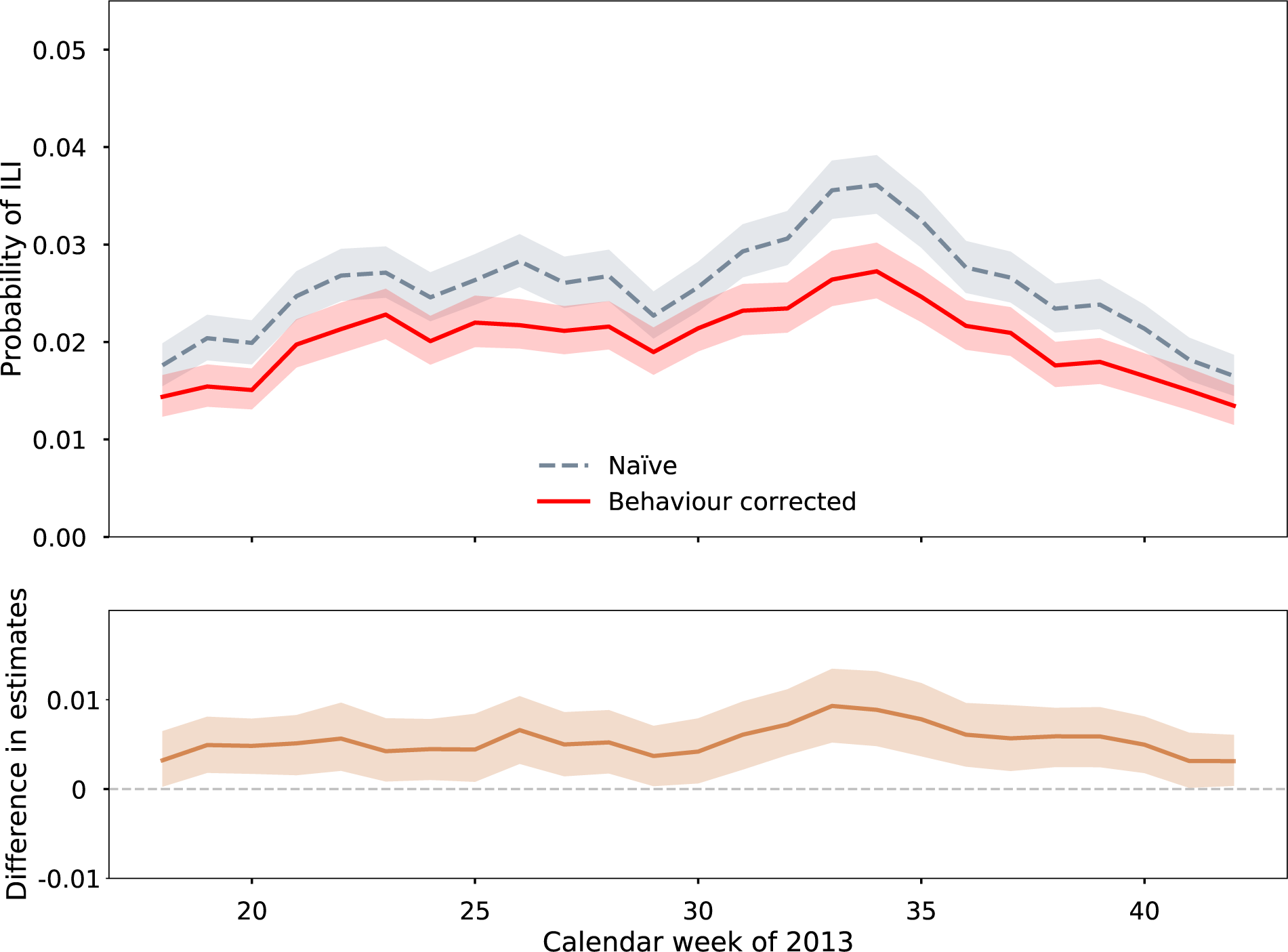
Comparison of model posterior estimate of the prevalence rate of ILI in the population to the naïve estimate for the winter of 2013, and the difference between the two distributions over time. Lines represent the median of the distribution and shaded regions are 95% credible intervals of the posterior distributions.

**Figure S.6:**
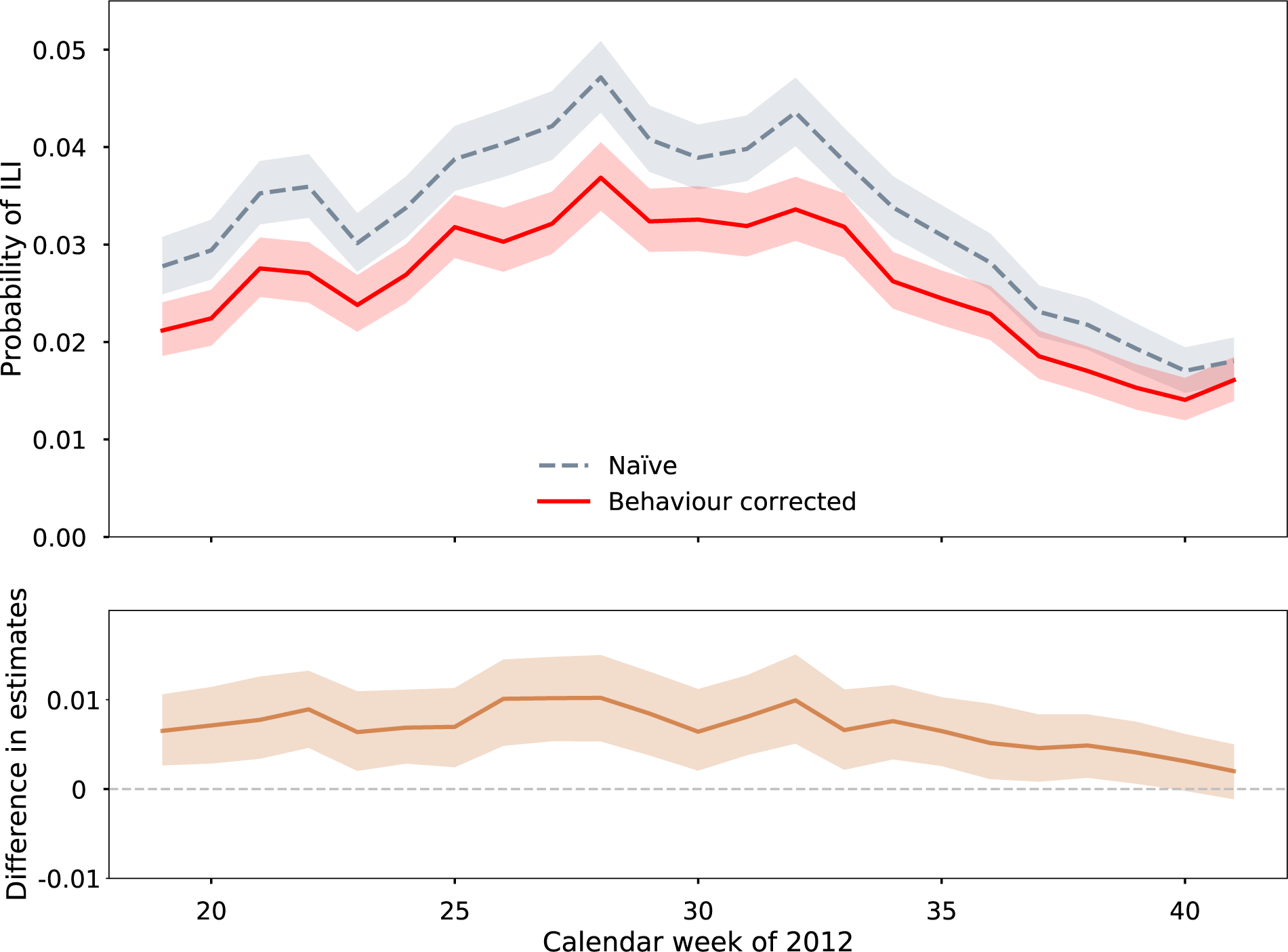
Comparison of model posterior estimate of the prevalence rate of ILI in the population to the naïve estimate for the winter of 2012, and the difference between the two distributions over time. Lines represent the median of the distribution and shaded regions are 95% credible intervals of the posterior distributions.

**Figure S.7:**
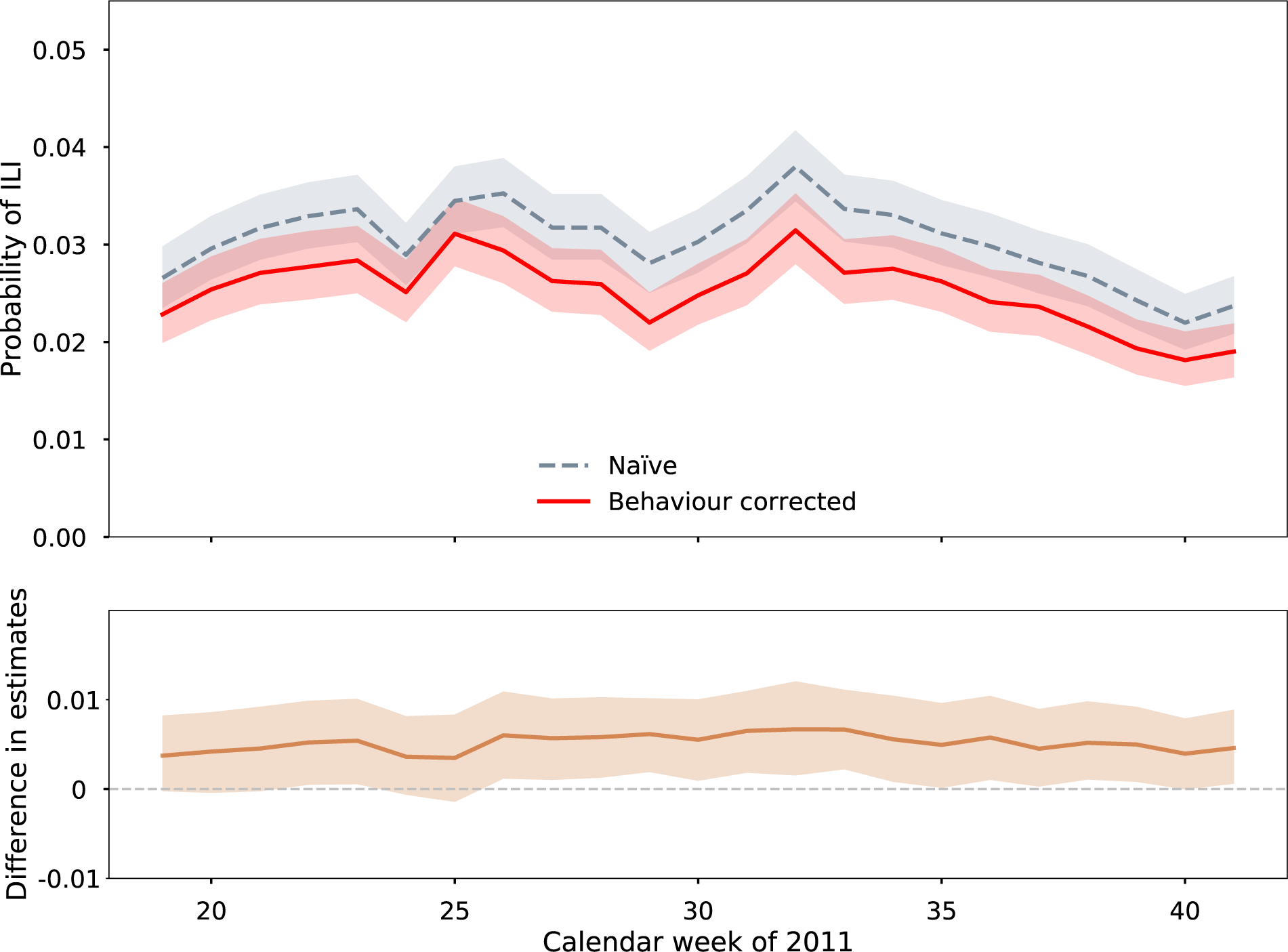
Comparison of model posterior estimate of the prevalence rate of ILI in the population to the naïve estimate for the winter of 2011, and the difference between the two distributions over time. Lines represent the median of the distribution and shaded regions are 95% credible intervals of the posterior distributions.

**Figure S.8:**
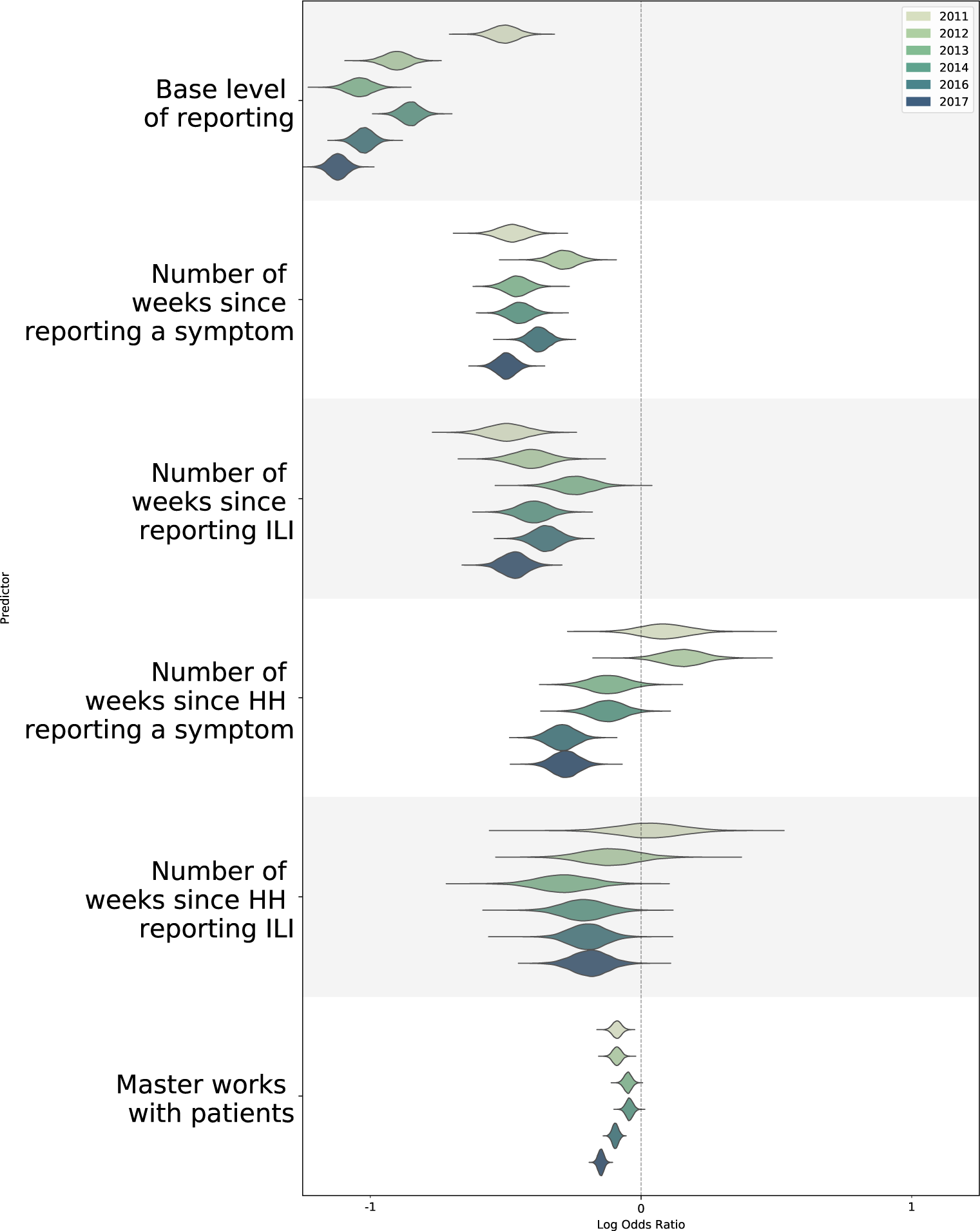
The marginal posterior densities of the log odds ratio of the first half of non-chronological regression coefficients for predicting the probability of an individual reporting for all years.

**Figure S.9:**
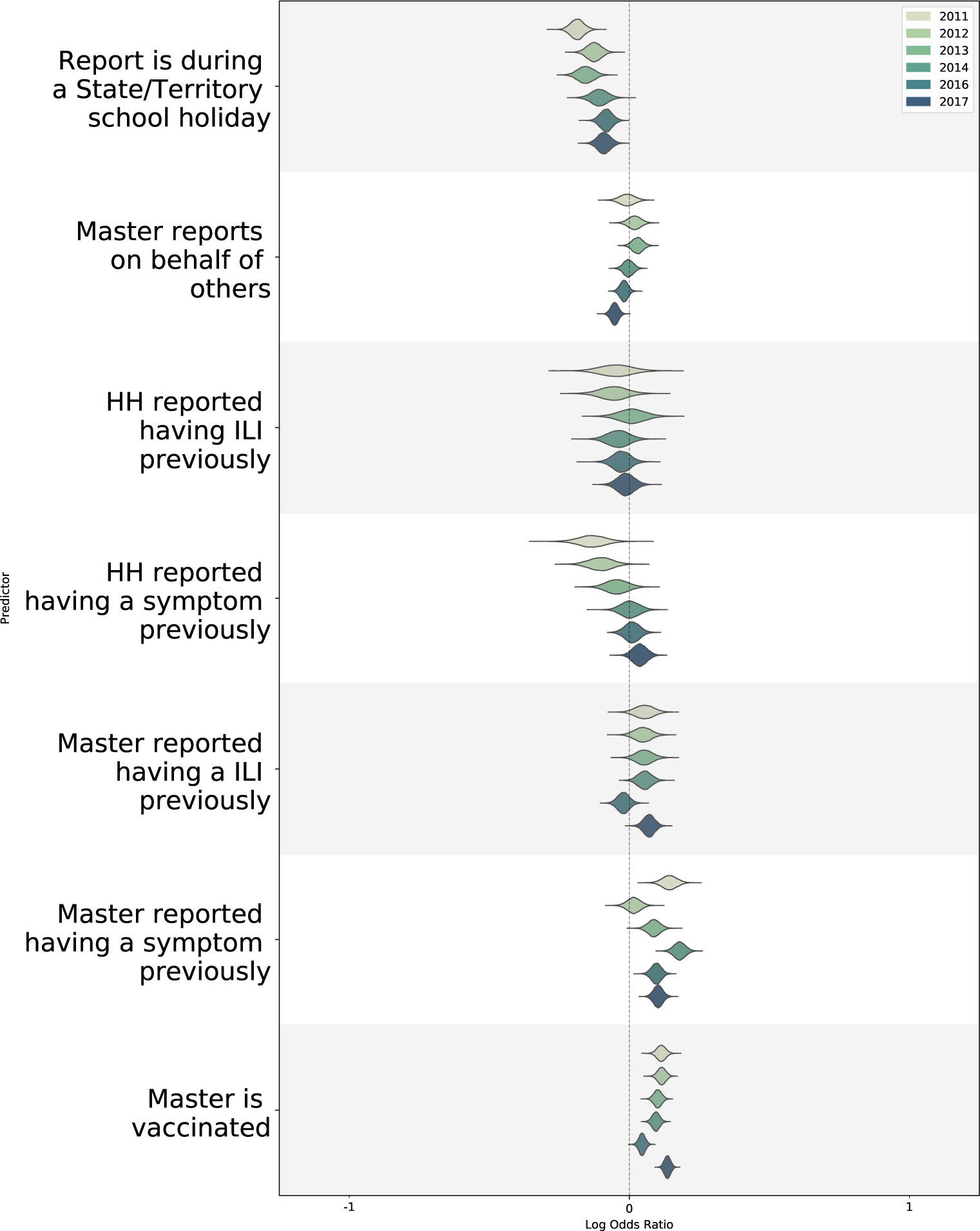
The marginal posterior densities of the log odds ratio of the second half of non-chronological regression coefficients for predicting the probability of an individual reporting for all years.

**Figure S.10:**
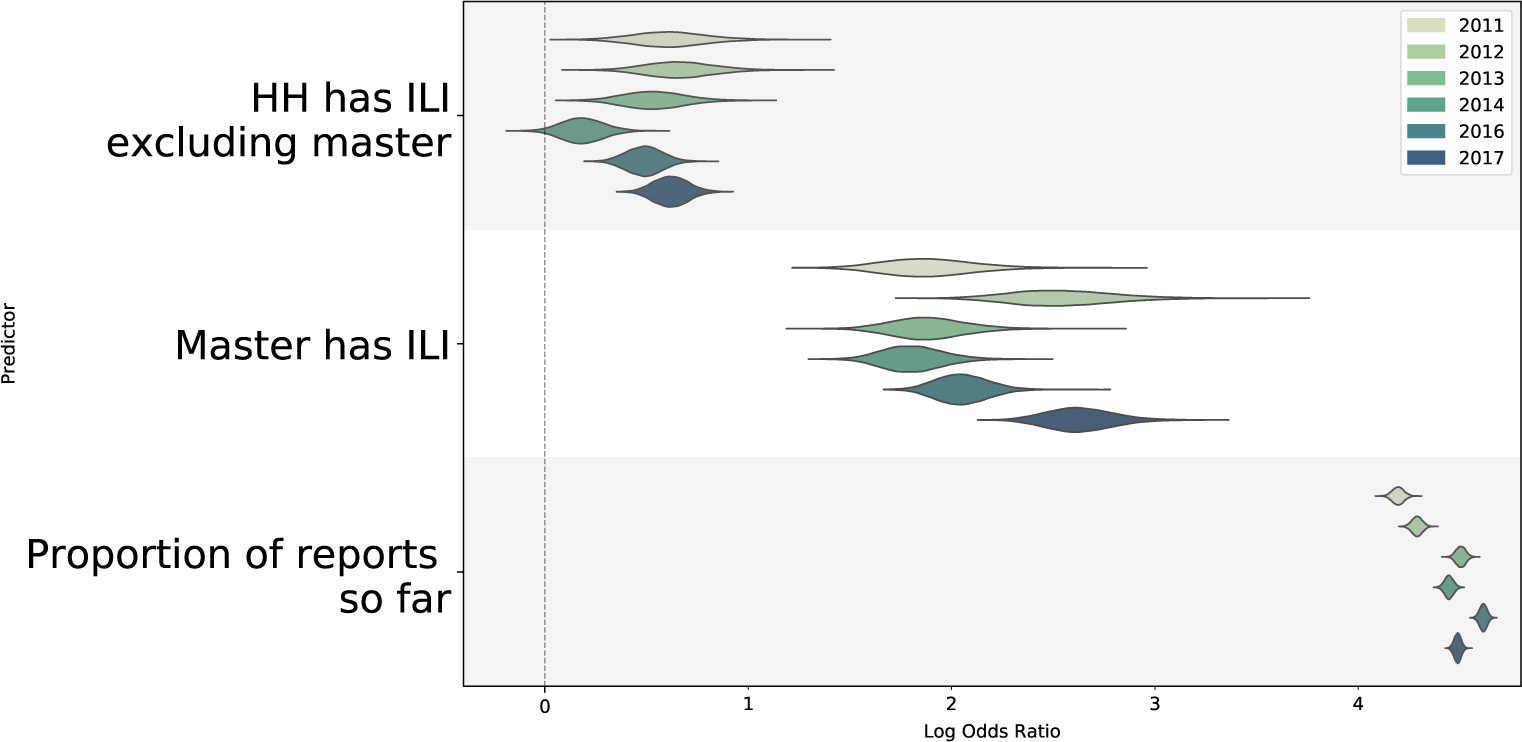
The marginal posterior densities of the log odds ratio of the largest regression coefficients for predicting the probability of an individual reporting for all years. Presented here are coefficients for if a member of the household has ILI and the proportion of reports submitted on-time

**Figure S.11:**
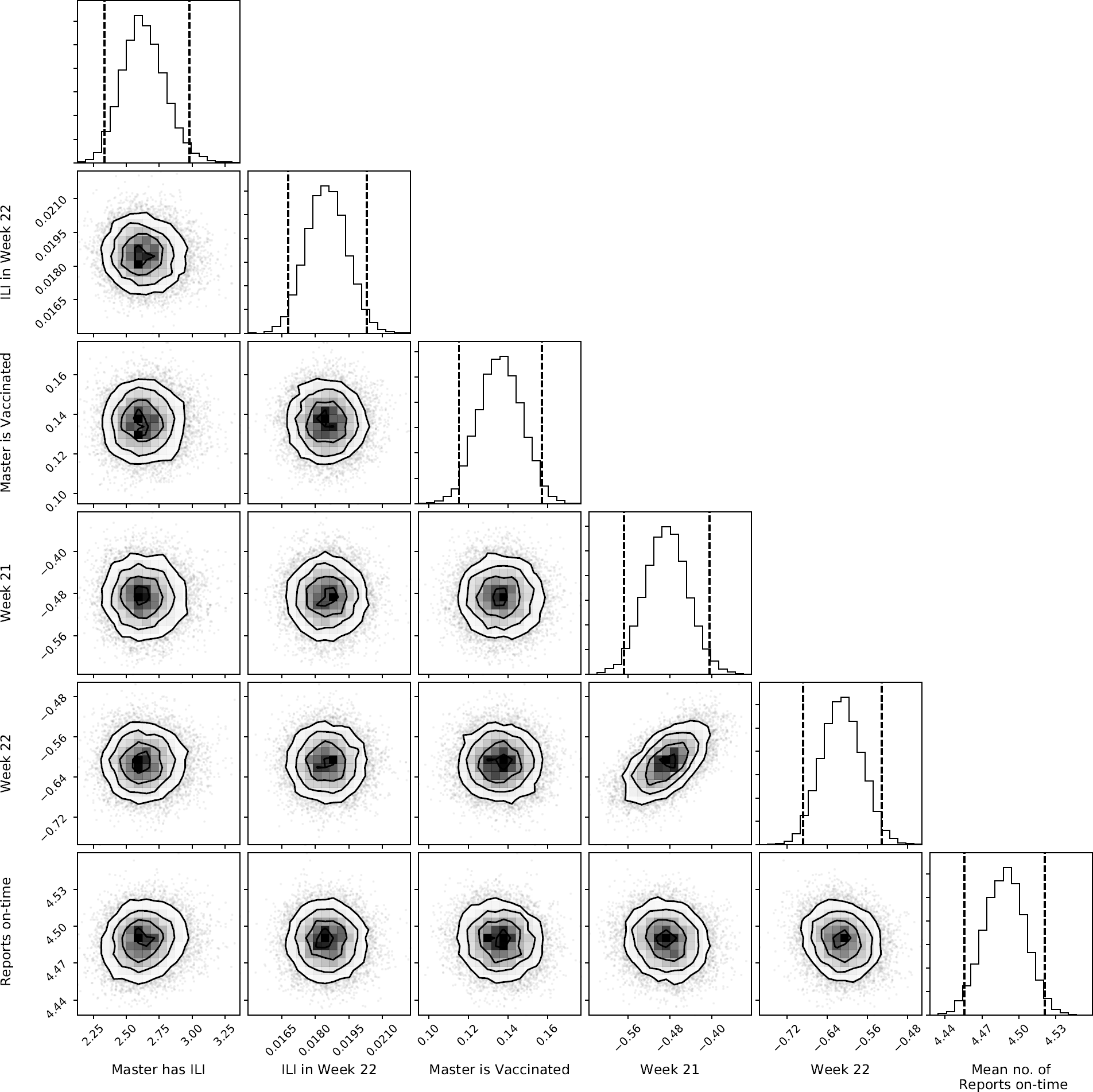
The bivariate kernel density of samples from the posterior for some parameters inferred from the 2017 data. Bivariate plots show little correlation between parameters, with some correlation in the chronological regression coefficients (Week 21 and Week 22).

**Figure S.12:**
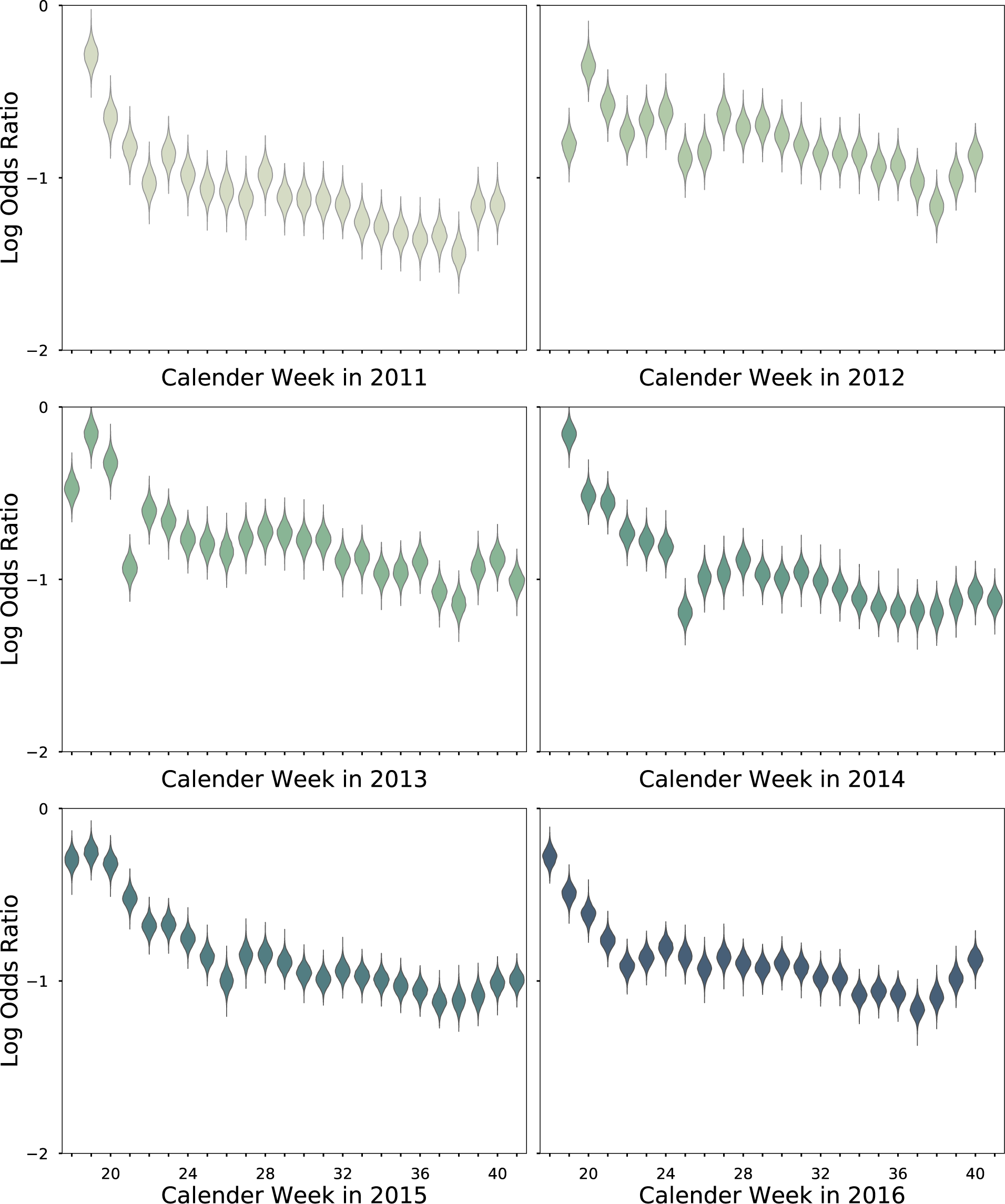
The marginal posterior densities of the log odds ratio of chronological regression coefficients for predicting the probability of an individual reporting in a given week for years 2011 to 2016.

**Figure S.13:**
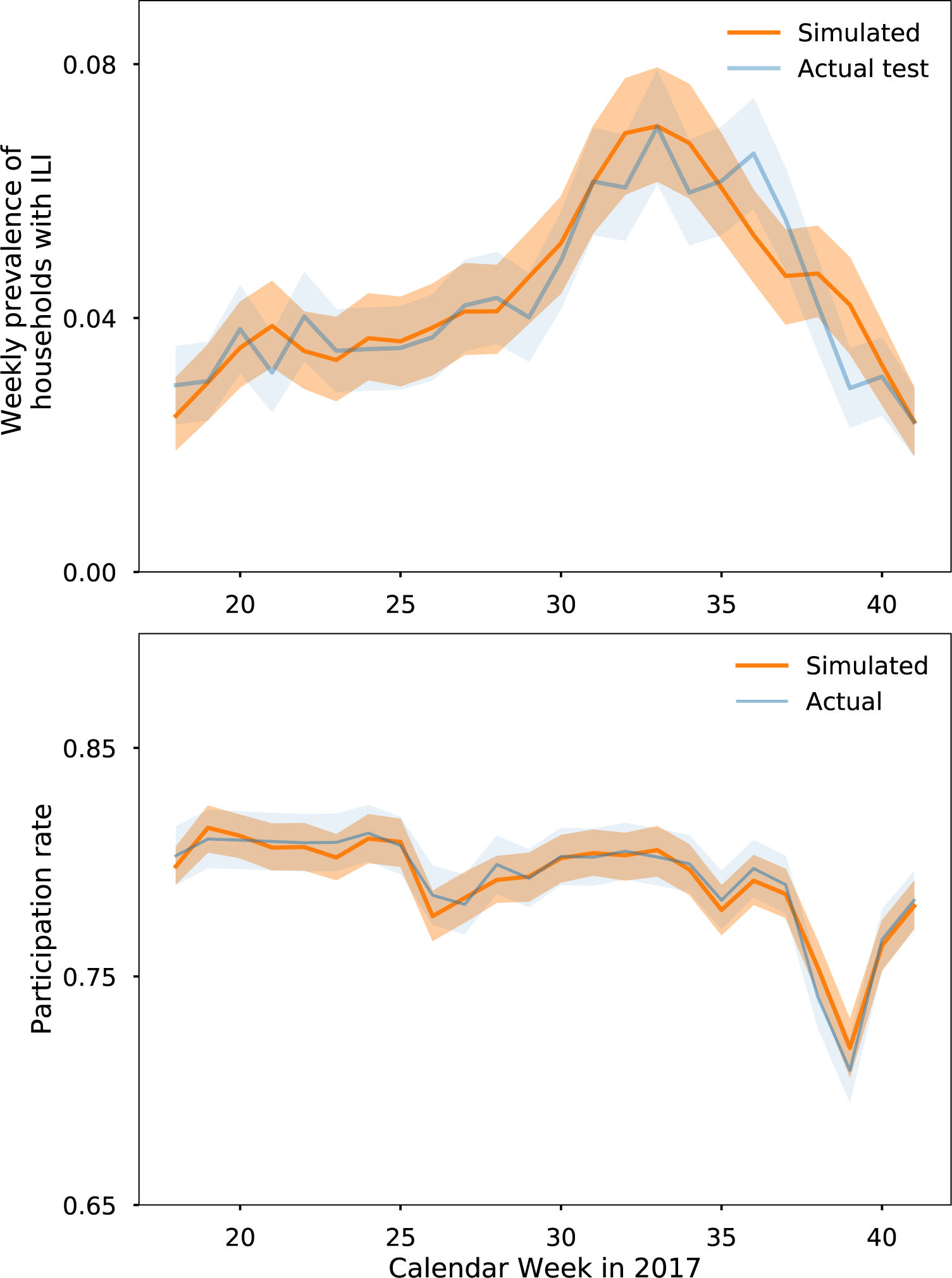
Cross validation of model predictions with actual outcomes of test set for the 2017 season. Lines represent the median and shaded regions the 95% credible intervals. The model is able to generate the data observed in the test set with high probability.

**Figure S.14:**
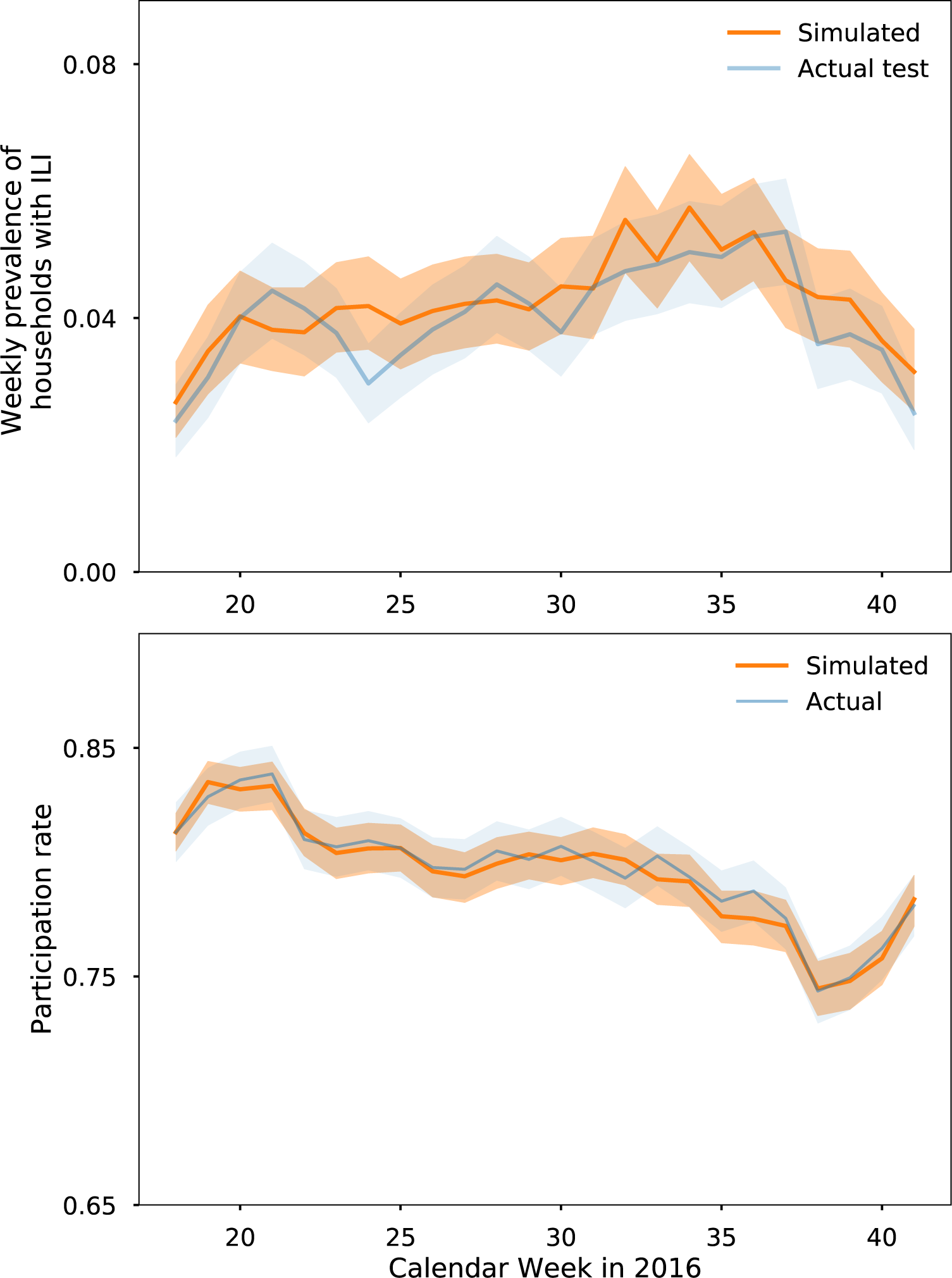
Cross validation of model predictions with actual outcomes of test set for the 2016 season. Lines represent the median and shaded regions the 95% credible intervals. The model is able to generate the data observed in the test set with high probability.

**Figure S.15:**
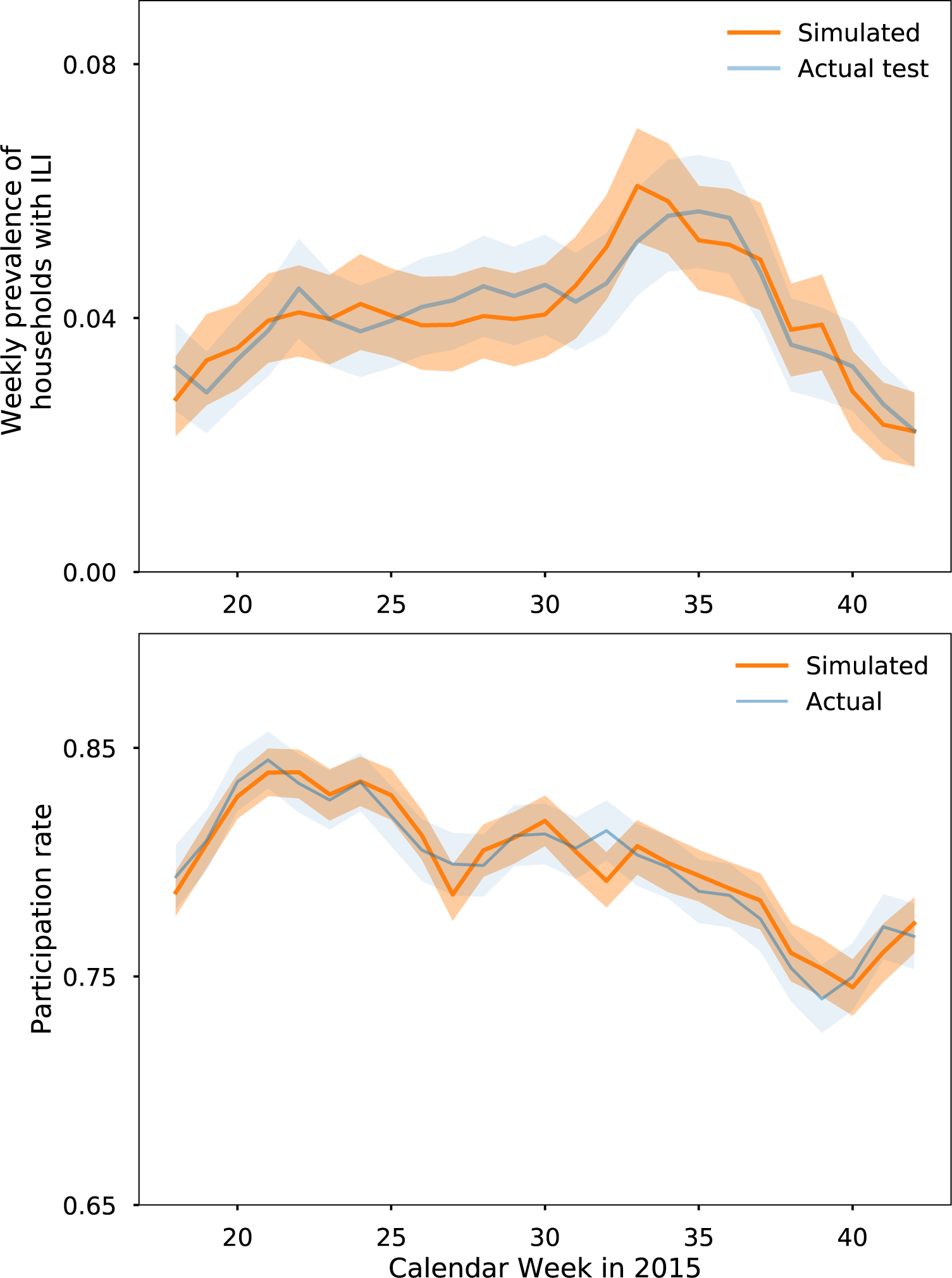
Cross validation of model predictions with actual outcomes of test set for the 2015 season. Lines represent the median and shaded regions the 95% credible intervals. The model is able to generate the data observed in the test set with high probability.

**Figure S.16:**
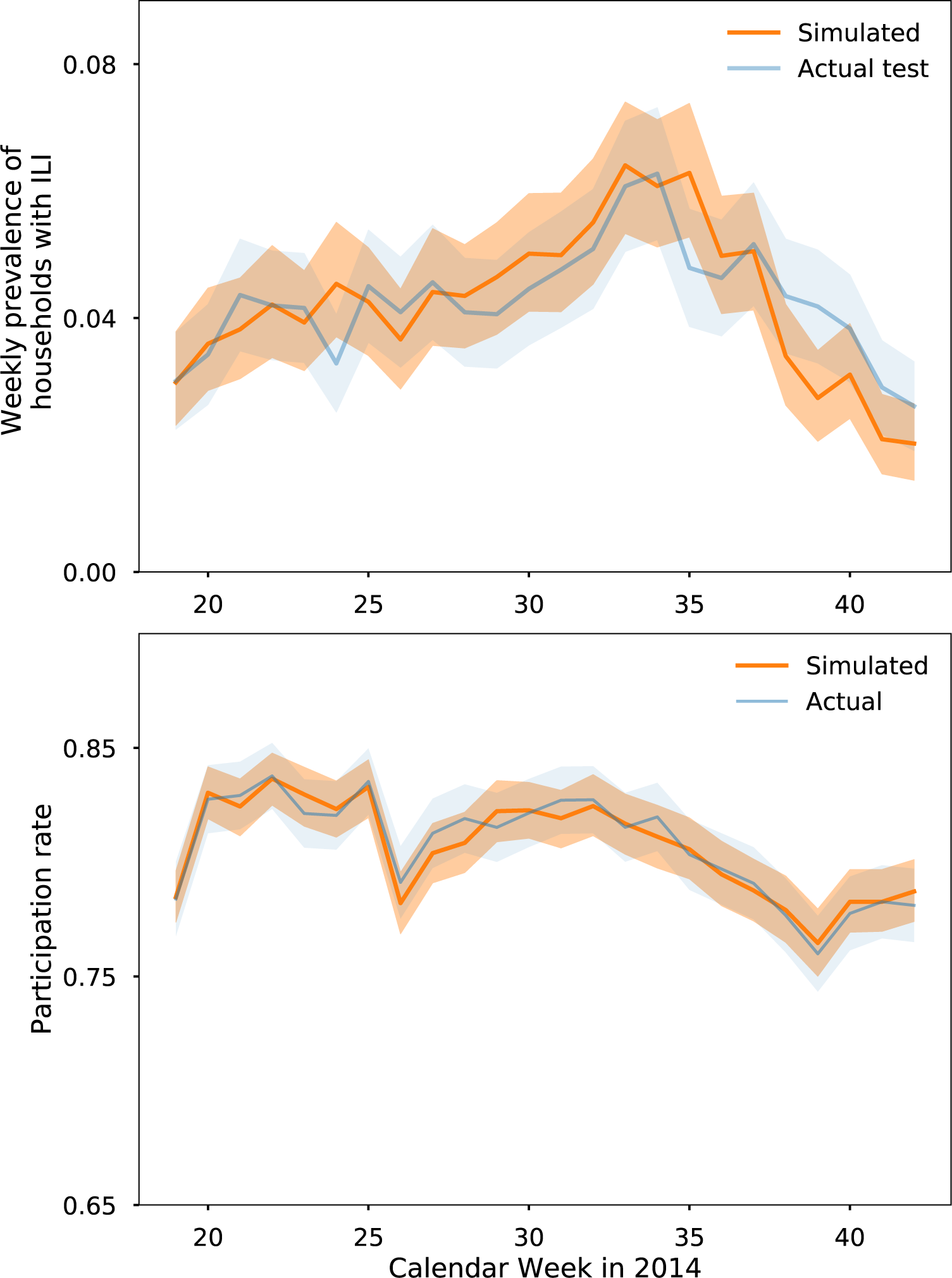
Cross validation of model predictions with actual outcomes of test set for the 2014 season. Lines represent the median and shaded regions the 95% credible intervals. The model is able to generate the data observed in the test set with high probability.

**Figure S.17:**
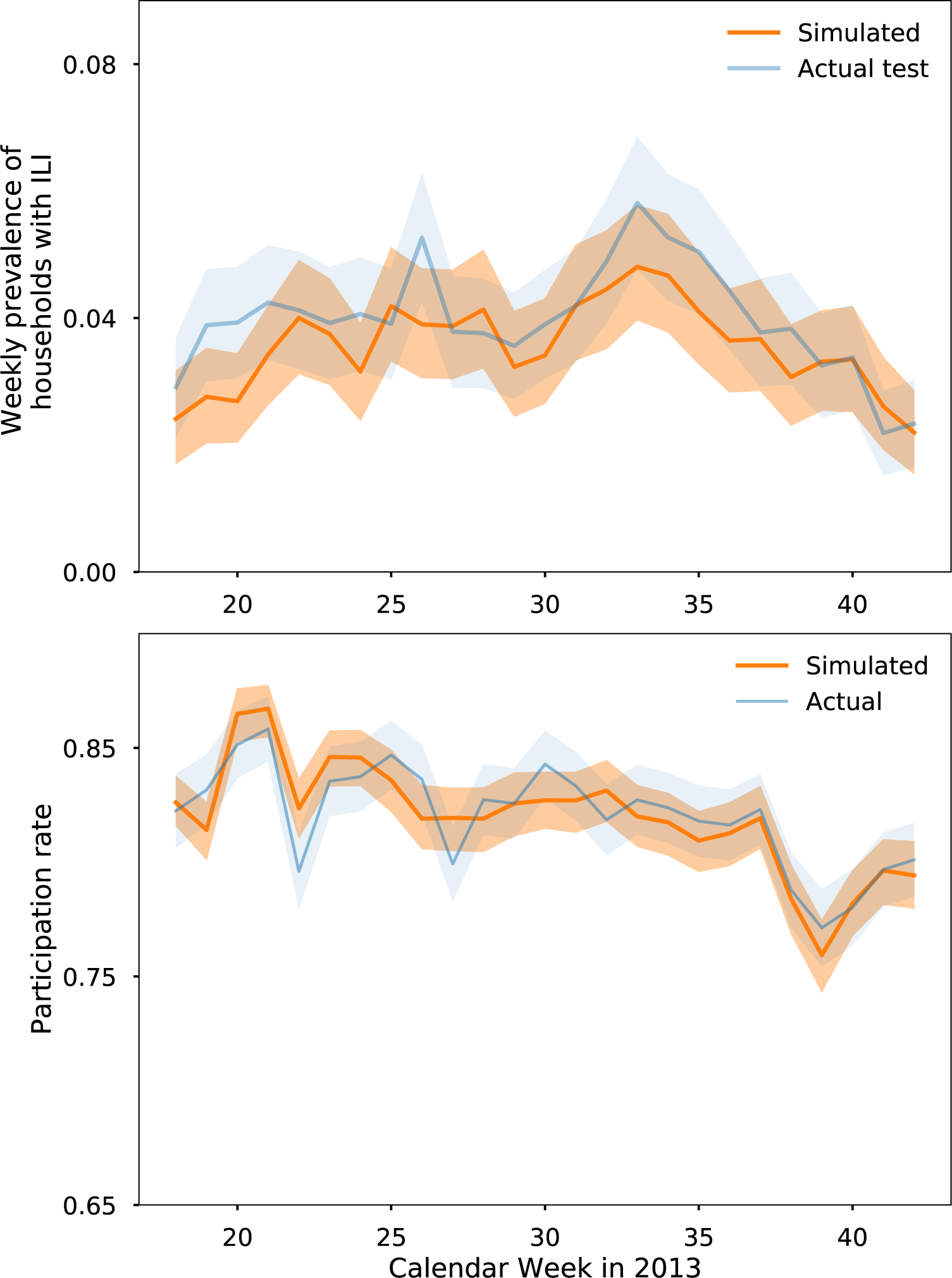
Cross validation of model predictions with actual outcomes of test set for the 2013 season. Lines represent the median and shaded regions the 95% credible intervals. The model is able to generate the data observed in the test set with high probability.

**Figure S.18:**
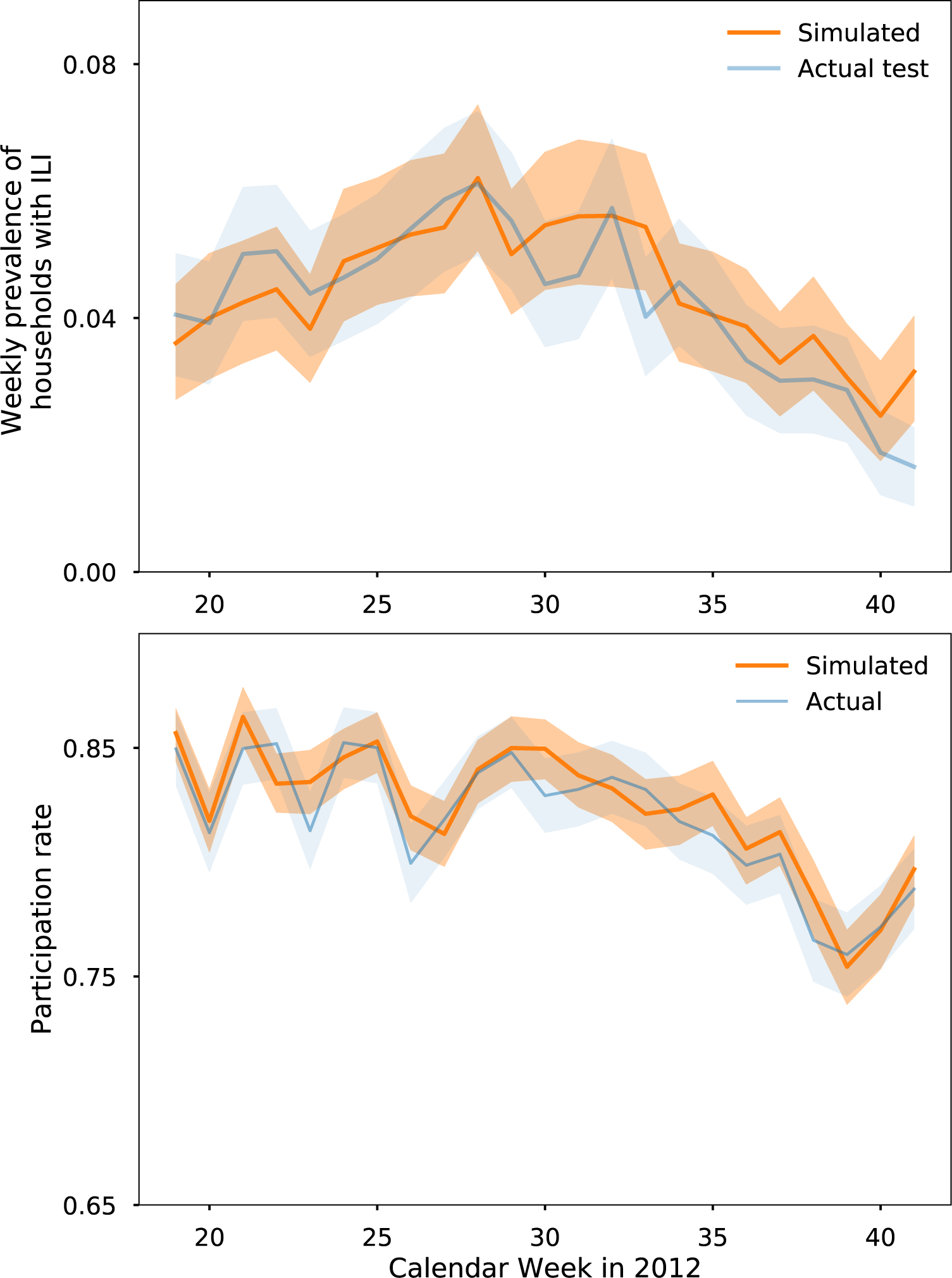
Cross validation of model predictions with actual outcomes of test set for the 2012 season. Lines represent the median and shaded regions the 95% credible intervals. The model is able to generate the data observed in the test set with high probability.

**Figure S.19:**
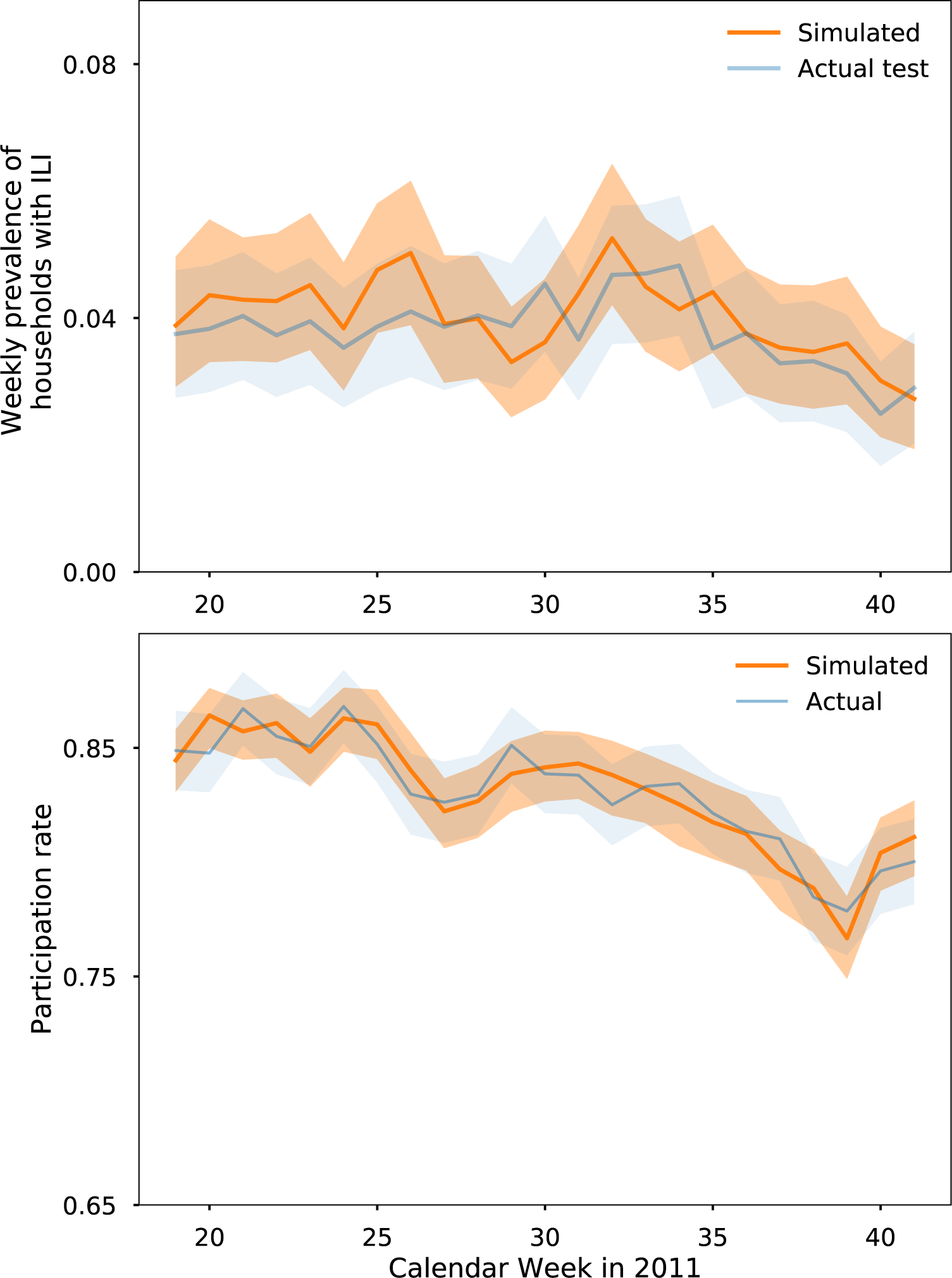
Cross validation of model predictions with actual outcomes of test set for the 2011 season. Lines represent the median and shaded regions the 95% credible intervals. The model is able to generate the data observed in the test set with high probability.

